# Placebo response in chronic peripheral neuropathic pain trials: systematic review and meta-analysis

**DOI:** 10.1101/2022.02.18.22271196

**Authors:** Gonçalo S Duarte, Beatrice Mainoli, Filipe B Rodrigues, Fábio Rato, Tiago Machado, Joaquim J Ferreira, João Costa

## Abstract

**Objective:** To estimate the magnitude of the placebo and nocebo responses in chronic peripheral neuropathic pain (CNP) and explore possible associations with trial characteristics.

**Methods:** We searched CENTRAL, MEDLINE, and Embase for randomized controlled trials (RCTs) from inception to May 2020. We included placebo-controlled RCTs of ≥8 weeks investigating first-line pharmacological interventions for CNP. Primary endpoints were the placebo response, the proportion of patients receiving placebo with pain intensity reduction (PIR) ≥30% from baseline, and the nocebo response, the proportion of patients receiving placebo experiencing adverse events (AEs). Screening, data extraction, and bias assessment (with the Cochrane risk of bias tool) were conducted by independent reviewers. We pooled data using a random-effects model.

**Results:** We included 50 trials, with a combined 5,693 participants allocated to placebo, conducted between 1998 and 2020. Overall, 38% of patients receiving placebo reported PIR≥30% (95% CI 34 to 42, I^2^=86%); 23% reported PIR≥50% (95% CI 20 to 26; I^2^=81%). 50% of patients receiving placebo reported AEs (95% CI 0.43 to 0.58; I^2^=97%); 2% reported serious AEs (95% CI 2 to 3; I^2^=58%). In patients receiving active interventions, the placebo response accounts for 75% of the treatment effect on PIR≥30%, and the nocebo response accounts for 75% of the AEs. Interpreted inversely, only 25% of responses and 25% of adverse events can be attributed to the intervention. Publication year positively correlated with PIR≥30% and negatively correlated with AEs. Female sex negatively correlated with AEs.

**Conclusions:** The placebo and nocebo responses in parallel-designed RCTs in CNP are substantial and should be considered in trial interpretation and in the design of future trials.

## INTRODUCTION

The placebo effect is a therapeutic benefit secondary to taking a pharmacologically inert substance,[4; 26]. Less commonly considered and understood, the nocebo effect is the emergence of adverse events secondary to the administration of a placebo.[3; 26]

The practical impact of these effects is highly relevant for the interpretation of trial results. A strong placebo effect has the tendency to decrease the difference between an active intervention compared with placebo, while a strong nocebo effect can be interpreted as a sign of disease progression,[8] and may mask potentially important safety signs. Thus, these phenomena can confound the assessment of both efficacy and safety of a drug in a clinical trial.[3; 13]

The International Association for the Study of Pain (IASP) definition of chronic peripheral neuropathic pain (CPNP) is “chronic pain caused by a lesion or disease of the peripheral somatosensory nervous system”.[28] The treatment of CPNP is challenging, with conventional analgesics typically being ineffective.[9] In fact, clinical practice guidelines do not recommend conventional analgesics as first-line treatment of CPNP, preferring drugs such as duloxetine, amitriptyline, topical capsaicin 8%, pregabalin, and gabapentin.[1; 12; 22; 23]

Previous meta-analyses have assessed the placebo and nocebo responses, though not exclusively in the chronic form of neuropathic pain.[6] As CPNP is notoriously more difficult to treat effectively compared with non-chronic neuropathic pain, and has a characteristically non-linear disease progression, the placebo and nocebo responses are particularly important to assess.[31] Although to assess the actual placebo effect would require trials to have a no-treatment arm, this is often not done in pain trials, in part due to ethical considerations.[19] Given this limitation, we deemed a measurement of the placebo response, the overall response of patients treated with placebo, to be the best alternative to the placebo effect, the response of patients treated with placebo minus the response of patients treated with no intervention.

In this systematic review and meta-analysis, we aim to estimate the placebo and nocebo responses in patients enrolled in randomized placebo-controlled trials assessing first-line interventions recommended for chronic peripheral neuropathic pain.

## METHODS

This systematic review is reported according to the PRISMA guidelines[21].

### Inclusion criteria

We considered any of the following interventions: single application of high□concentration (8%) topical capsaicin, tricyclic antidepressants, serotonin-norepinephrine reuptake inhibitors (SNRIs), or calcium channel alpha-2-delta ligands. The reason for selecting these drug classes as interventions to be studied is because most clinical guidelines (including the International Association for the Study of Pain,[12] the past European Federation of Neurological Societies,[1] the National Institute for Health and Care Excellence,[23] and the Canadian Pain Society[22]) currently provide strong recommendations for using these drugs as first-line therapy.

We included randomized controlled trials (RCTs) comparing any of the aforementioned drug classes versus placebo or an active comparator. In order to avoid possible bias due to population enrichment, we excluded trials which excluded non-responders after an active run-in period.[10] Regarding study duration, as there is evidence of over-estimating treatment effects in shorter trials,[15] we excluded trials with a duration of fewer than 8 weeks. We included trials enrolling adults (i.e., ≥ 18 years) with peripheral neuropathic pain in the context of chemotherapy-induced peripheral neuropathy (CIPN), HIV□neuropathy (HIVN), painful diabetic peripheral neuropathy (PDPN), postherpetic neuralgia (PHN), mixed populations, and post-surgical neuropathy. These data were analyzed in the pooled population across all conditions.

Studies had to report quantitative data on at least one of the following endpoints:

#### Primary endpoints

The placebo response, defined as the proportion of participants with participant□reported pain intensity reduction (PIR) of 30% or greater in the placebo arm; the nocebo response, defined as the proportion of participants experiencing adverse events (AEs) in the placebo arm.

#### Secondary endpoints

PIR of 50% or greater; Patient Global Impression of Change (PGIC) much or very much improved; global pain assessment (such as with a visual analogue scale (VAS), numerical rating scale (NRS), or other); discontinuations due to lack of efficacy and adverse events (AEs); participants with serious adverse event (SAE); and specific AEs.

All endpoints were binary in nature, and therefore events refer to the number of patients experiencing the endpoint under analysis (e.g., the number of participants with PIR of 30% or greater). The percentage cutoffs of 30% and 50% for PIR were chosen as these are suggested by the Cochrane pain group.[7] We also extracted the same data from the intervention arms, to establish a point of comparison. No language, year of publication, or publication status restrictions were applied.

### Information sources

We searched MEDLINE, Embase, and CENTRAL databases, the WHO International Clinical Trials Registry Platform, and clinicaltrials.gov, from inception to May 2020.

Two reviewers (BM, GSD) independently screened the search results. Disagreements were resolved by consensus. The reasons for exclusion were recorded at the full-text screening stage. Three reviewers (BM, GSD, TM) extracted study data following a pre-established data collection form. Data from studies’ plots were retrieved through Plot Digitizer version 2.6.8. When studies presented different estimates of the endpoint of interest, we extracted the most precise or adjusted measures.

Risk of bias was independently evaluated by three authors (BM, GSD, TM) using the Cochrane risk of bias tool, where seven domains were qualitatively classified as at high, unclear, or low risk of bias.[18] The overall risk of bias for each RCT was divided as high or low risk, with high risk being those RCTs in which at least one domain was assessed at a high risk of bias, or more than three domains were had a rating of unclear.[10]

### Summary measures

Data were derived from the last measured within-group response in the placebo or intervention arms of RCTs. Whenever possible, we retrieved and analyzed intention-to-treat data. Regarding the PIR endpoints, the methods used for determining the PIR were different between trials, and we used the proportions as reported. We used R for statistical analysis and to derive forest plots. We used Freeman-Tukey transformed proportions[16] in a random-effects model to pool data due to the anticipated heterogeneity among the included trials.[29] Between-study variance was estimated using the Paule-Mandel method.[24] We reported pooled dichotomous data reporting 95% confidence intervals (CIs). Statistical heterogeneity between trial results was assessed using I^2^ and tau^2^.[17]

### Additional analyses

Assuming that the treatment and placebo/nocebo responses are additive and not interactive, we conducted a further analysis. The proportion of symptoms not attributable to the pharmacological action (PSN) of a drug was also established, [2; 11; 20] using the following formula:

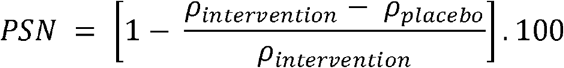

ρ_*intervention*_: pooled proportion of an event in the intervention arm, ρ_*placebo*_: pooled proportion of the same event in the placebo arm.

We conducted meta-regressions for the primary trial endpoints to test for the impact of possible confounders. We opted not to conduct multiple meta-regressions due to the limited number of trials. We tested the impact of transdermal application, pain duration at baseline, trial duration, overall trial risk of bias, the underlying condition, percentage of female participants, median participant age, and year of publication. To reduce the likelihood of overfitting, before building our model we tested for multicollinearity (see supplementary material).

## RESULTS

The results of the search are shown in Figure 1. We included 50 RCTs with 5,693 participants allocated to placebo (15,524 participants overall), published between 1998 and 2020 (Table 1). Trial references and characteristics are summarized in the supplementary material. The median trial sample size was 308 (interquartile range [IQR] 167 to 400). The median participant age was 60 (IQR 53 to 65), and 39.7% of the overall trial participants were women (6,158 of 15,524). Forty-eight trials (96%) used a double-blind design. The primary trial endpoints were assessed after eight weeks in 21 trials (42%), 10-12 weeks in 24 trials (48%), and longer in 5 trials (10%). Only one trial was not published.

**Figure 1.**
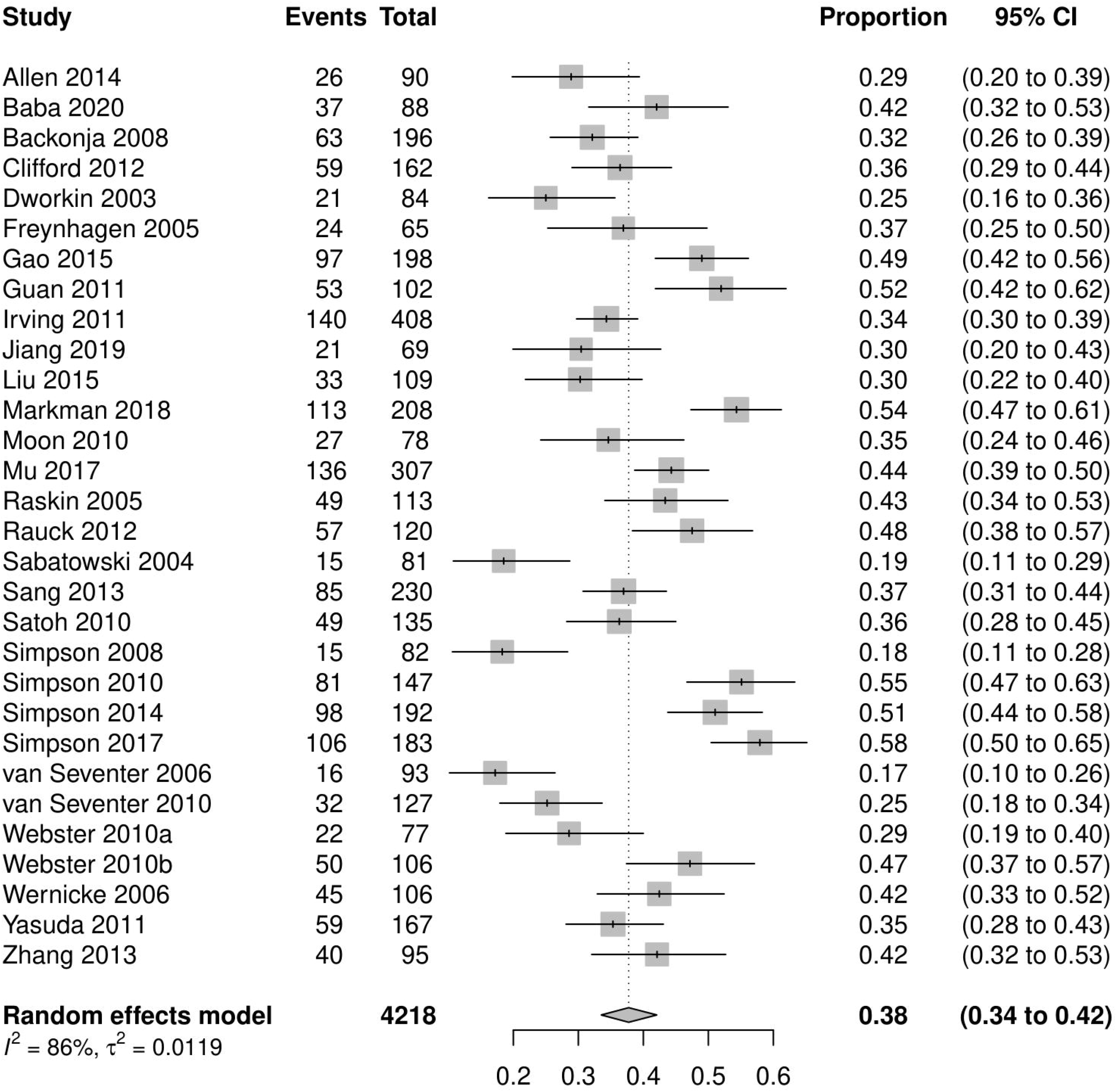
PRISMA flow chart.

**Table 1.** Main results for pooled analyses.

| Endpoint | Trials | Patients receiving placebo |  |  |  | Patients receiving active interventions |  |  |  | Proportion of symptoms non-pharmacological (%) |
| --- | --- | --- | --- | --- | --- | --- | --- | --- | --- | --- |
|  |  | Events | Total participants | Proportion (95% CI) | I<sup>2</sup> | Events | Total participants | Proportion (95% CI) | I<sup>2</sup> |  |
| PIR≥30%<br>(Placebo response) | 30 | 1669 | 4218 | 38 (34 to 42) | 86 | 2537 | 4925 | 51 (47 to 56) | 89 | 75 |
| AEs<br>(Nocebo response) | 37 | 2316 | 4649 | 50 (43 to 58) | 97 | 3912 | 6040 | 67 (60 to 74) | 97 | 75 |
| PIR≥50% | 32 | 1052 | 4257 | 23 (20 to 27) | 81 | 2411 | 6605 | 36 (33 to 39) | 82 | 64 |
| PGIC much or very much improved | 27 | 977 | 3224 | 26 (21 to 33) | 93 | 1955 | 4587 | 42 (37 to 46) | 88 | 62 |
| Discontinuations due to AE | 43 | 255 | 5246 | 4 (3 to 6) | 77 | 774 | 7887 | 8 (6 to 11) | 92 | 50 |
| Discontinuations due to lack of efficacy | 37 | 203 | 4442 | 4 (3 to 6) | 80 | 172 | 6631 | 2 (1 to 2) | 80 | - |
| Serious AEs | 39 | 135 | 4983 | 2 (2 to 3) | 58 | 193 | 7041 | 2 (2 to 3) | 68 | - |

AE, adverse event. CI, confidence interval. PGIC, patient global impression of change. PIR, pain intensity reduction.

Notably, 18 trials were conducted in patients with PDPN (2,142 participants allocated to placebo), 14 trials were conducted in patients with PHN (1,822 participants allocated to placebo), 11 trials were conducted in patients with more than one condition (974 participants allocated to placebo), six trials were conducted in patients with HIVN (686 participants allocated to placebo), and one trial was conducted in patients with CIPN (69 participants allocated to placebo).

The drug classes of the active intervention arms were gabapentin and/or pregabalin (31 trials, 62%), SNRIs (seven trials, 14%), transdermal capsaicin 8% (seven trials, 14%), and tricyclic antidepressants (five trials, 10%).

### Risk of bias

Regarding the overall risk of bias, 30 trials (60%) had a low risk of bias, and 20 trials (40%) had a high risk of bias. Industry funded 40 trials (80%). The risk of bias in individual trials is available in the supplementary material.

### Data synthesis

#### Placebo response

Based on 1,669 events, 38% of patients receiving placebo reported PIR≥30% (95% CI 34 to 42; I^2^ = 86%; 30 trials; n = 4,218; Figure 2). Based on 4,925 events, 51% of patients receiving active interventions reported PIR≥30% (95% CI 47 to 56; I^2^ = 89%; n = 4,925). The calculated PSN is 75%.

**Figure 2.**
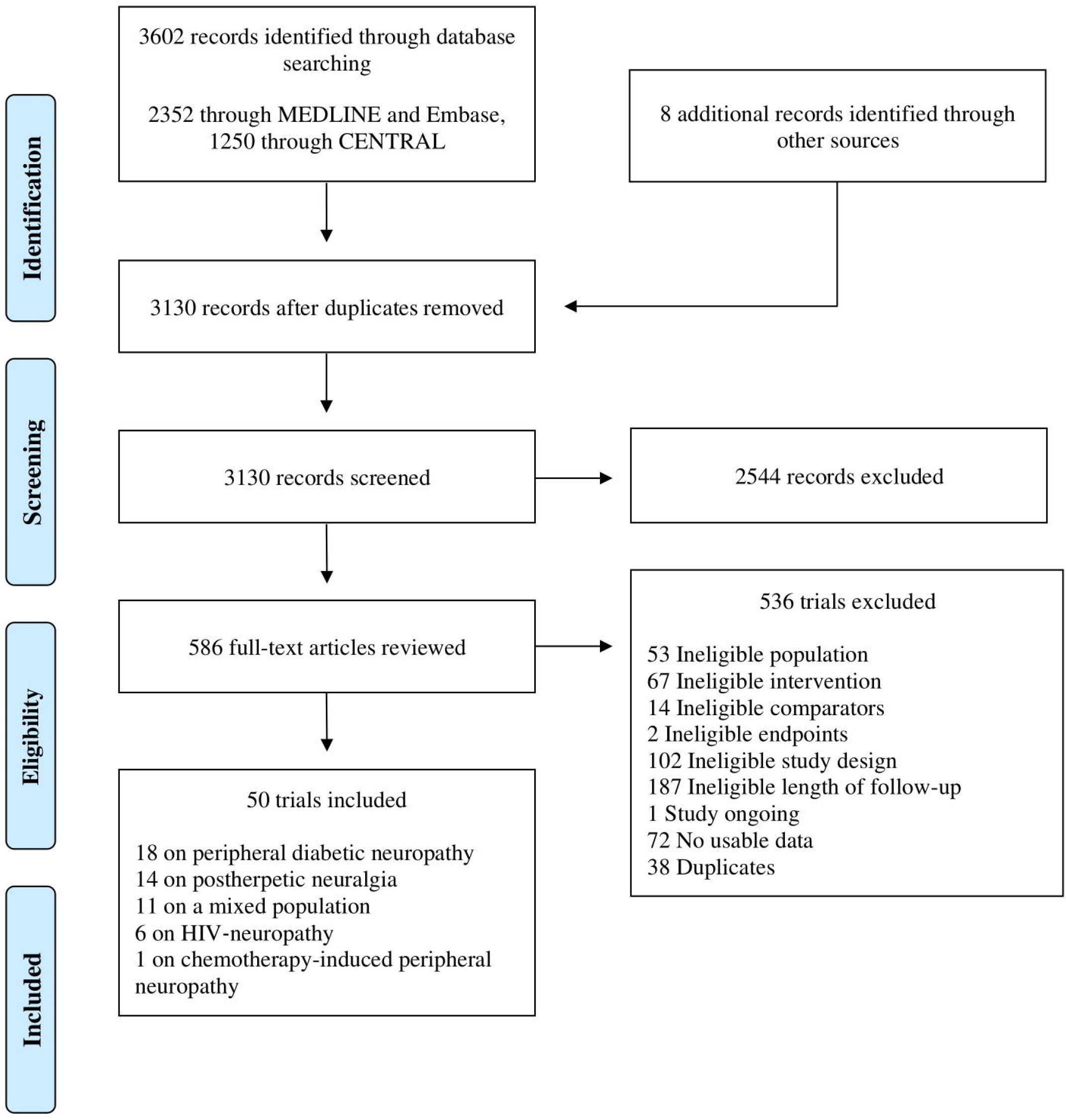
Patients receiving placebo with pain intensity reduction (PIR) ≥30%.

#### Nocebo response

Based on 2,316 events, 50% of patients receiving placebo reported AEs (95% CI 43 to 58; I^2^ = 97%; 37 trials; n = 4,649; Figure 3). Based on 3,912 events, 67% of patients receiving active interventions reported AEs (95% CI 60 to 74; I^2^ = 97; n = 6,040). The calculated PSN is 75%.

**Figure 3.**
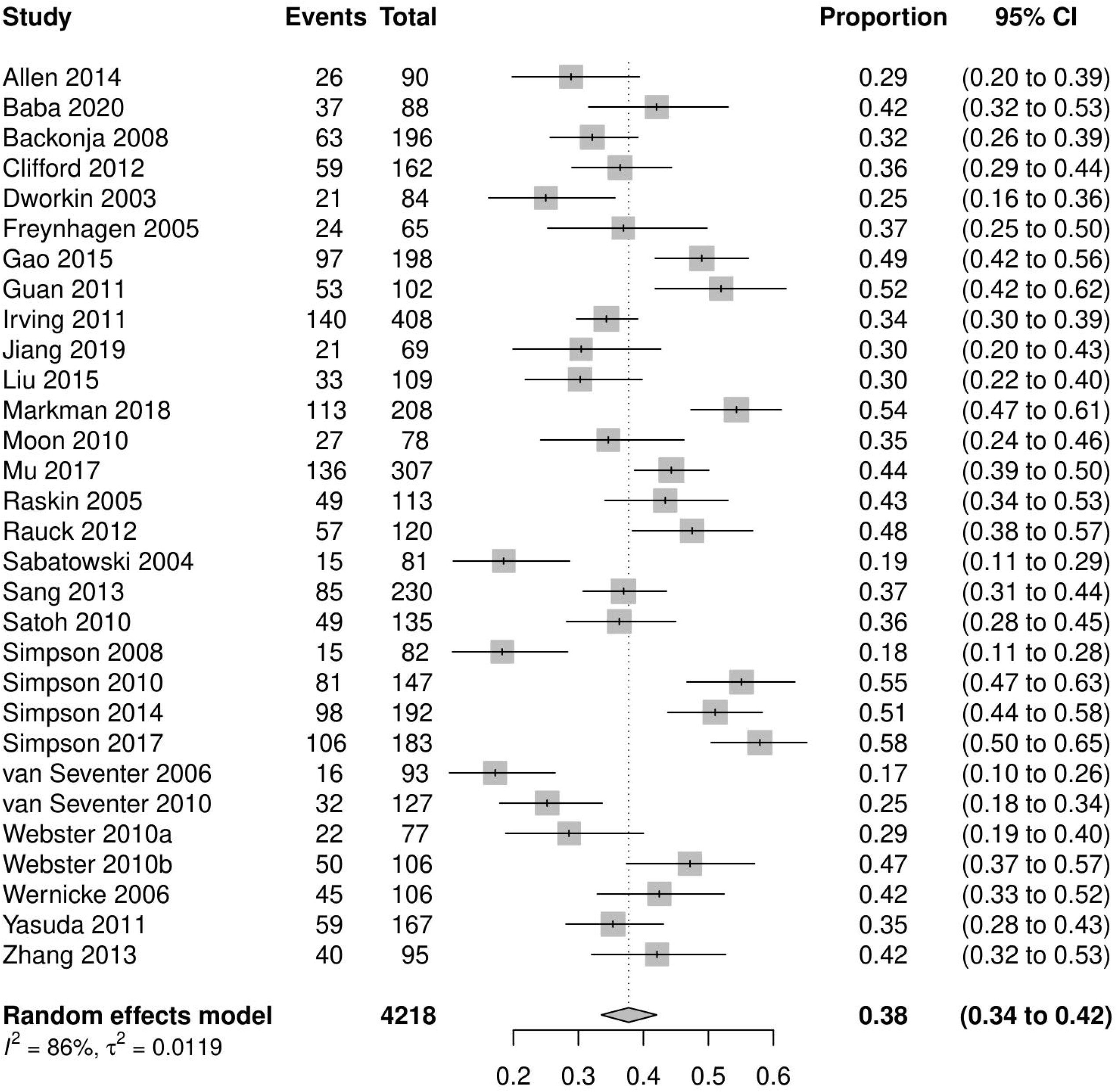
Patients receiving placebo with adverse events (AEs).

#### Secondary endpoints

All pooled results can be seen in Table 1. 23% of patients receiving placebo reported PIR≥50% (95% CI 20 to 27; I^2^ = 81%). 26% of patients receiving placebo reported PGIC much or very much improved (95% CI 21 to 33; I^2^ = 93%). 4% of patients receiving placebo discontinued due to AEs (95% CI 3 to 6; I^2^ = 77%). 4% of patients receiving placebo discontinued due to lack of efficacy (95% CI 3% to 6%; I^2^ = 80%). 2% of patients receiving placebo reported SAEs (95% CI 2% to 3%; I^2^ = 58%).

The calculated PSN was 64% for PIR≥50%, 62% for PGIC much or very much improved, 50% for discontinuations due to AEs, and 100% for SAEs.

#### Meta-regressions

Table 2 reports the results of the meta-regressions. We found that year of publication was positively associated with PIR≥30% (p = 0.0067) and negatively correlated with AEs (p = 0.0371). We also found a negative association with AEs and the percentage of female trial participants (p = 0.00061), though AEs was positively correlated with trial duration (p = 0.0447).

**Table 2.**
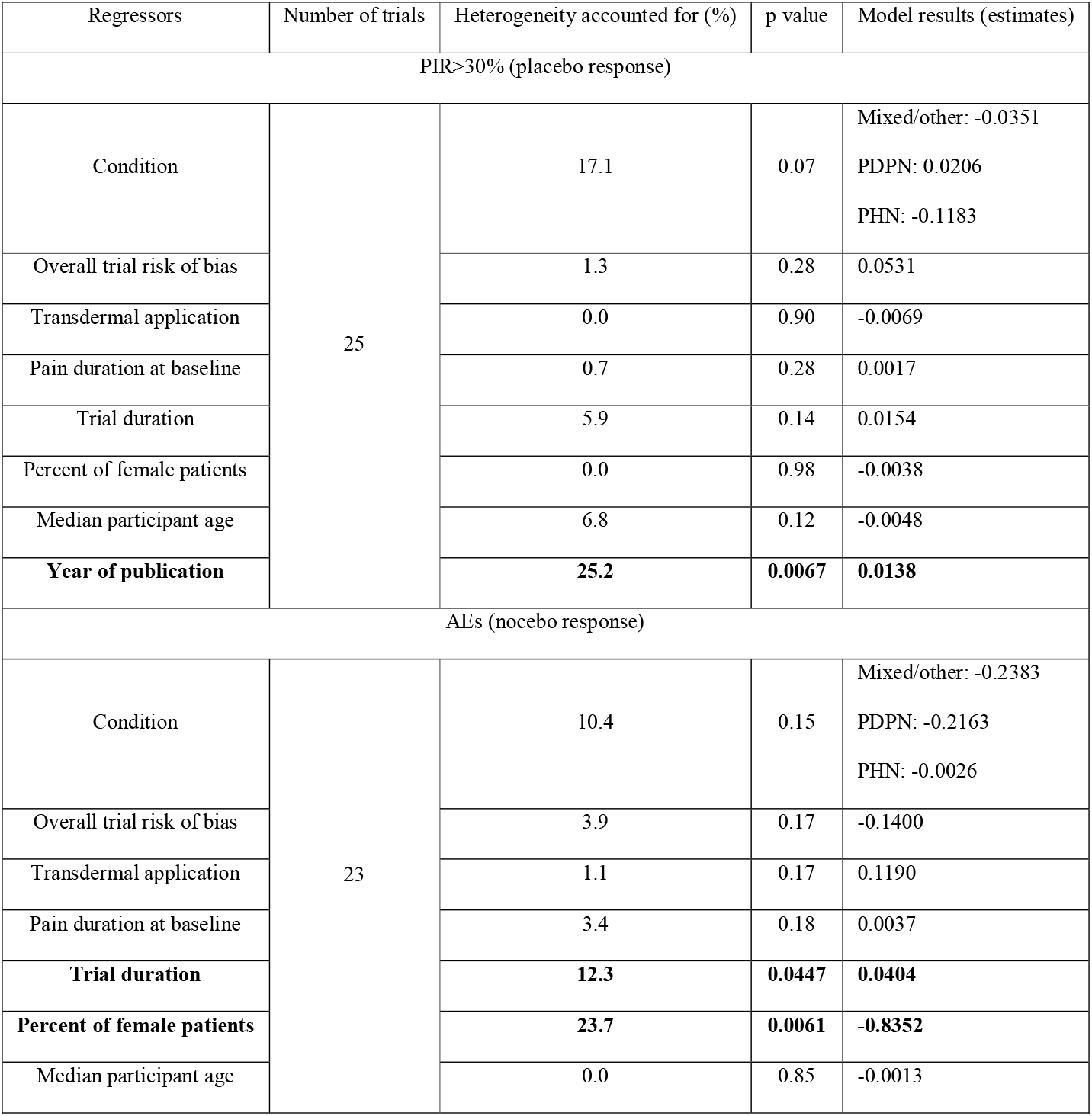
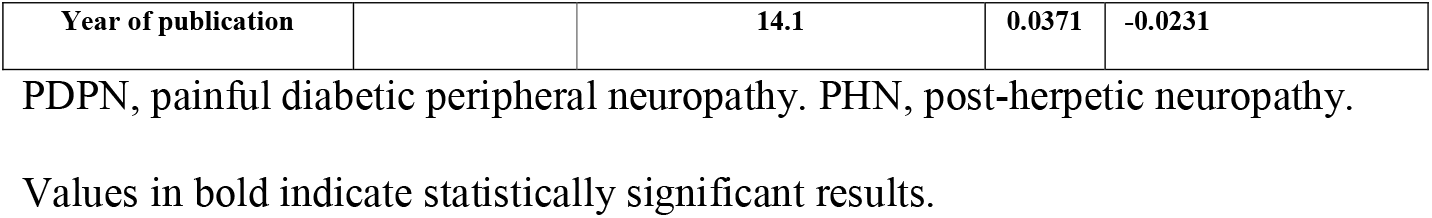
Results of meta-regressions.

| Regressors | Number of trials | Heterogeneity accounted for (%) | p value | Model results (estimates) |
| --- | --- | --- | --- | --- |
| PIR≥30% (placebo response) |  |  |  |  |
| Condition | 25 | 17.1 | 0.07 | Mixed/other: -0.0351<br>PDPN: 0.0206<br>PHN: -0.1183 |
| Overall trial risk of bias |  | 1.3 | 0.28 | 0.0531 |
| Transdermal application |  | 0.0 | 0.90 | -0.0069 |
| Pain duration at baseline |  | 0.7 | 0.28 | 0.0017 |
| Trial duration |  | 5.9 | 0.14 | 0.0154 |
| Percent of female patients |  | 0.0 | 0.98 | -0.0038 |
| Median participant age |  | 6.8 | 0.12 | -0.0048 |
| Year of publication |  | 25.2 | 0.0067 | 0.0138 |
| AEs (nocebo response) |  |  |  |  |
| Condition | 23 | 10.4 | 0.15 | Mixed/other: -0.2383<br>PDPN: -0.2163<br>PHN: -0.0026 |
| Overall trial risk of bias |  | 3.9 | 0.17 | -0.1400 |
| Transdermal application |  | 1.1 | 0.17 | 0.1190 |
| Pain duration at baseline |  | 3.4 | 0.18 | 0.0037 |
| Trial duration |  | 12.3 | 0.0447 | 0.0404 |
| Percent of female patients |  | 23.7 | 0.0061 | -0.8352 |
| Median participant age |  | 0.0 | 0.85 | -0.0013 |
| Year of publication |  | **14.1** | **0.0371** | **-0.0231** |

PDPN, painful diabetic peripheral neuropathy. PHN, post-herpetic neuropathy.
Values in bold indicate statistically significant results.

## DISCUSSION

Our meta-analysis demonstrated that a significant number of patients with chronic peripheral neuropathic pain report significant improvements across multiple endpoints, constituting evidence of a strong positive placebo effect. However, half of the analyzed patients report having suffered AEs, constituting evidence of a strong nocebo effect. That both these phenomena were observed from a theoretically inert intervention is interesting.

To our knowledge, this is the first systematic review addressing the placebo effect in chronic neuropathic pain. Previous reviews assessed the placebo and nocebo responses in non-chronic and not exclusively peripheral neuropathic pain, showing considerable effect.[6] However, our pooled estimate of the nocebo response is similar to that estimated by Papadopoulos and colleagues, namely 52% (95% CI, 36 to 68). Likewise, in other forms of pain such as

As calculated by the PSN, 75% of the AEs among patients treated with an active intervention can be attributed to placebo. This result suggests that most AEs is not causally related to the medication but can be seen across patients with chronic peripheral neuropathic pain, independently of their medication., although the Hawthorne effect cannot be excluded.

More interestingly perhaps, 75% of the patients that reported PIR≥30% can have that effect attributable to placebo or Hawthorne effects. Additionally, this effect was 64% for PIR≥50% and 52% for PGIC much or very much improved. This is significant for clinicians who must interpret the likely causal relationship between an event and the medication. It is also relevant for the choice of endpoints in future trials of CNP, since PGIC much or very much improved is likely the best choice to reduce the impact of the placebo effect, and so will more easily demonstrate differences between placebo and active interventions.[14]

Regarding the generalizability of these data, in conditions such as chronic pain, the doctor-patient relationship likely influences the number and readiness with which patients report adverse events. This is likely stronger in the context of clinical trials, as can be seen by the very low rates of treatment discontinuation due to both AEs and due to lack of efficacy.

It is important to compare the observed results to previous research on placebo. Regarding nocebo effect, our results are in line with other studies in neurological diseases, namely restless legs syndrome, where an estimated 45% of patients receiving placebo reported AEs[30] and Parkinson’s disease, where an estimated 56% of patients receiving placebo reported AEs.[20] Regarding cervical dystonia, a neurologic condition associated with chronic pain, an estimated 53% of patients receiving placebo reported AEs.[11]

Although these results were obtained from the analysis of placebo arms only, factors other than the placebo and nocebo effect may influence these results. The two most important factors to consider are the Hawthorne Effect,[19] whereby patients under observation will experience a larger response, and regression to the mean. The Hawthorne Effect has been studied in acute induced pain, where people treated with open-label placebo referred a 21% lower median pain rating when compared with no treatment.[27] In the context of neuropathic pain, the regression to the mean may be of particular importance owing to the highly variable and non-linear natural history of the disease.[31] Thus, oscillations in disease status over time can be interpreted as an improvement or worsening of the disease, despite being part of the normal disease fluctuation.

Pain is a highly subjective outcome without external objective methods to validate a patient’s self-reported assessment, and so may be more easily modulated by external factors. The underlying conditions that cause the chronic neuropathic pain are also of importance. These conditions can influence the course of the pain a patient feels, and may also contribute to manifestations that, in the context of clinical trials, can be interpreted as AEs. However, pain relief after placebo administration has been associated with decreased activity in pain processing regions.[32] Studies have shown involvement of expectations and emotional feelings in placebo effect across chronic pain conditions, including neuropathic pain.[25] There is evidence of a similar effect regarding nocebo.[5]

Our meta-regressions showed statistically significant results for year of publication, which was positively correlated with PIR≥30% and negatively correlated with AEs. This is of interest as it suggests that over time trials in pain research may be

## CONCLUSIONS

Placebo is not an inert intervention, having not only beneficial effects but also potentially significant detrimental consequences. The high risk of AEs among placebo-treated patients suggests that current best evidence on safety and tolerability is possibly confounded by the nocebo response; at the same time, the magnitude of placebo response calculated in this context indicates its substantial implication in the assessment of efficacy endpoints. Our data suggest that the placebo response is smaller when efficacy is assessed using PGIC much or very much improved, compared with PIR≥30% or PIR≥50%. Year of publication was associated with a larger placebo response and a smaller nocebo response. Female sex was negatively associated with reported AEs. Our data further corroborate the power of expectation and the emotional component in the management of chronic peripheral neuropathic pain, be it positive or negative.

## Data Availability

All data produced in the present work are contained in the manuscript

## AUTHOR ROLES

1. Research project:
  A. Conception: GSD
  B. Organization: GSD
  C. Execution: BM, GSD, JC, TM
2. Statistical Analysis:
  A. Design: BM, GSD
  B. Execution: GSD
  C. Review and Critique: BM, FR, JC, JJF, TM
3. Manuscript Preparation:
  A. Writing of the first draft: BM
  B. Review and Critique: FR, GSD, JC, JJF, TM

All authors have approved the final article

