## Supplementary material for "Placebo response in chronic peripheral neuropathic pain trials: systematic review and meta-analysis"

Laboratório de Farmacologia Clínica e Terapêutica

Faculdade de Medicina de Lisboa, Av. Prof. Egas Moniz, 1649-028 Lisboa, Portugal

**Phone number:** (+351) 21 797 34 53; **Fax number:** (+351) 21 781 96 88.

### Search strategies

#### CENTRAL search strategy

1. MeSH descriptor: [Capsaicin] explode all trees
2. (capsaicin OR capsaicine OR capsici OR axsain OR capsidol OR capsig OR capsin OR capsina OR capsiplast OR capzasin-P OR dolorac OR gelcen OR katrum OR “No pain-HP” OR priltam OR “R-gel” OR zacin OR zostrix OR capsicum)
3. MeSH descriptor: [Administration, topical] explode all trees
4. (topical* OR cutaneous OR dermal OR transcutaneous OR transdermal OR percutaneous OR skin OR massage OR embrocation OR gel OR ointment OR aerosol OR cream OR crème OR lotion OR foam OR liniment OR spray OR rub OR balm OR salve OR emulsion OR oil OR patch OR plaster)
5. (#1 OR #2) AND (#3 OR #4)
6. MeSH descriptor: [Antidepressive Agents, Tricyclic] explode all trees
7. (amitriptyline hydrochloride OR amoxapine OR clomipramine hydrochloride OR dosulepin hydrochloride OR dothiepin hydrochloride OR doxepin OR imipramine hydrochloride OR lofepramine OR nortriptyline OR trimipramine OR tricyclic* OR desipramine OR florpiramine OR dibenzepin OR iprindole OR protriptyline OR amoxapine OR opipramol)
8. #6 OR #7
9. MeSH descriptor: [Anticonvulsants] explode all trees
10. (gabapentin OR pregabalin OR neurontin OR lyrica OR gabapentinoids)
11. MeSH descriptor: [Serotonin Uptake Inhibitors] explode all trees
12. (SSRI* OR SNRI*)
13. (serotonin* AND (reuptake OR re‐uptake) AND inhibitor*)
14. (citalopram OR dapoxetin* OR escitalopram OR fluoxetin* OR fluvoxamin* OR paroxetin* OR sertralin* OR desvenlafaxin* OR duloxetin* OR milnacipran OR venlafaxin*)
15. #9 OR #10 OR #11 OR #12 OR #13 OR #14
16. MeSH descriptor: [Diabetic Neuropathies] explode all trees
17. MeSH descriptor: [Peripheral Nervous System Diseases] explode all trees
18. MeSH descriptor: [Neuralgia] explode all trees
19. (neuropath* OR diabet* post-herpetic OR neuralgia OR phantom OR stump)
20. #16 OR #17 OR #18 OR #19
21. (#5 OR #8 OR #15 OR #20) AND
22. Limit 21 to Trials

#### MEDLINE search strategy

1. exp Capsaicin/
2. (capsaicin OR capsaicine OR capsici OR axsain OR capsidol OR capsig OR capsin OR capsina OR capsiplast OR capzasin-P OR dolorac OR gelcen OR katrum OR “No pain-HP” OR priltam OR “R-gel” OR zacin OR zostrix OR capsicum).mp.
3. or/1-2
4. exp Administration, topical/
5. (topical* OR cutaneous OR dermal OR transcutaneous OR transdermal OR percutaneous OR skin OR massage OR embrocation OR gel OR ointment OR aerosol OR cream OR creme OR lotion OR foam OR liniment OR spray OR rub OR balm OR salve OR emulsion OR oil OR patch OR plaster).mp.
6. (1 or 2) and (4 or 5)
7. exp Antidepressive Agents, Tricyclic/
8. (tricyclic$ adj3 antidepres$).tw.
9. Amitriptyline/
10. amitriptyline hydrochloride.tw.
11. Amoxapine/
12. amoxapine.tw.
13. Clomipramine/
14. clomipramine hydrochloride.tw.
15. Dothiepin/
16. (dosulepin hydrochloride or dothiepin hydrochloride).tw.
17. Doxepin/
18. doxepin.tw.
19. Imipramine/
20. imipramine hydrochloride.tw.
21. Lofepramine/
22. lofepramine.tw.
23. Nortriptyline/
24. nortriptyline.tw.
25. Trimipramine/
26. trimipramine.tw.
27. tricyclic$.tw.
28. Desipramine/
29. desipramine.tw.
30. florpiramine.tw.
31. dibenzepin.tw.
32. Iprindole/
33. iprindole.tw.
34. Protriptyline/
35. protriptyline.tw.
36. or/7-35
37. exp Serotonin Uptake Inhibitors/
38. (SSRI* or SNRI*).mp.
39. (serotonin* and (reuptake or re‐uptake) and inhibitor*).mp.
40. (citalopram or dapoxetin* or escitalopram or fluoxetin* or fluvoxamin* or paroxetin* or sertralin* or desvenlafaxin* or duloxetin* or milnacipran or venlafaxin*).mp.
41. or/37-40
42. exp Anticonvulsants/ or exp gamma-Aminobutyric Acid/ or gabapentin.mp.
43. gaba agents.mp. or exp GABA Agents/
44. gabapentinoids.mp.
45. pregabalin.mp. or exp Pregabalin/
46. lyrica.mp.
47. neurontin.mp.
48. or/42-47
49. exp Diabetic neuropathies/
50. exp Peripheral Nervous System Diseases/
51. exp Neuralgia/
52. (neuropath* OR diabet* post-herpetic OR neuralgia OR phantom OR stump).mp.
53. or/49-52
54. "randomized controlled trial".pt.
55. (random$ or placebo$ or single blind$ or double blind$ or triple blind$).ti,ab.
56. (retraction of publication or retracted publication).pt.
57. or/54-56
58. (animals not humans).sh.
59. ((comment or editorial or meta-analysis or practice-guideline or review or letter or journal correspondence) not "randomized controlled trial").pt.
60. (random sampl$ or random digit$ or random effect$ or random survey or random regression).ti,ab. not "randomized controlled trial".pt.
61. or/58-60
62. 57 not 61
63. (6 or 36 or 41 or 48) and 53 and 62

#### Embase search strategy

1. exp Capsaicin/
2. (capsaicin OR capsaicine OR capsici OR axsain OR capsidol OR capsig OR capsin OR capsina OR capsiplast OR capzasin-P OR dolorac OR gelcen OR katrum OR “No pain-HP” OR priltam OR “R-gel” OR zacin OR zostrix OR capsicum).mp.
3. or/1-2
4. exp Administration, topical/
5. topical* OR cutaneous OR dermal OR transcutaneous OR transdermal OR percutaneous OR skin OR massage OR embrocation OR gel OR ointment OR aerosol OR cream OR creme OR lotion OR foam OR liniment OR spray OR rub OR balmOR salve OR emulsion OR oil OR patch OR plaster).mp.
6. (1 or 2) and (4 or 5)
7. exp tricyclic antidepressant agent/
8. Amitriptyline/
9. amitriptyline hydrochloride.tw.
10. Amoxapine/
11. amoxapine.tw.
12. Clomipramine/
13. clomipramine hydrochloride.tw.
14. Dothiepin/
15. (dosulepin hydrochloride or dothiepin hydrochloride).tw.
16. Doxepin/
17. doxepin.tw.
18. Imipramine/
19. imipramine hydrochloride.tw.
20. Lofepramine/
21. lofepramine.tw.
22. Nortriptyline/
23. nortriptyline.tw.
24. Trimipramine/
25. trimipramine.tw.
26. tricyclic$.tw.
27. Desipramine/
28. desipramine.tw.
29. florpiramine.tw.
30. dibenzepin.tw.
31. Iprindole/
32. iprindole.tw.
33. Protriptyline/
34. protriptyline.tw.
35. or/7-34
36. exp Serotonin Uptake Inhibitors/
37. (SSRI* or SNRI*).mp.
38. (serotonin* and (reuptake or re‐uptake) and inhibitor*).mp.
39. (citalopram or dapoxetin* or escitalopram or fluoxetin* or fluvoxamin* or paroxetin* or sertralin* or desvenlafaxin* or duloxetin* or milnacipran or venlafaxin*).mp.
40. or/36-39
41. exp gabapentin/ or gabapentin.mp.
42. anticonvulsants.mp. or exp anticonvulsive agent/
43. neurontin.mp.
44. gabapentinoids.mp.
45. pregabalin/
46. pregabalin.mp.
47. lyrica.mp.
48. or/41-47
49. exp Diabetic neuropathies/
50. exp Peripheral Nervous System Diseases/
51. exp Neuralgia/
52. or/49-51
53. (neuropath* OR diabet* post-herpetic OR neuralgia OR phantom OR stump).mp.
54. (random$ or placebo$ or single blind$ or double blind$ or triple blind$).ti,ab.
55. RETRACTED ARTICLE/
56. or/53-55
57. (animal$ not human$).sh,hw.
58. (book or conference paper or editorial or letter or review).pt. not exp randomized controlled trial/
59. (random sampl$ or random digit$ or random effect$ or random survey or random regression).ti,ab. not exp randomized controlled trial/
60. or/56-59
61. 56 not 60
62. (6 or 35 or 40 or 48) 52 and 61

### Reference list of included trials

| Trial | Reference |
| --- | --- |
| Allen 2014 | [1] |
| Arezzo 2008 | [2] |
| Baba 2020 | [3] |
| Backonja 1998 | [4] |
| Backonja 2008 | [5] |
| Clifford 2012 | [6] |
| Dworkin 2003 | [7] |
| Eerdekens 2016 | [8] |
| Eftekharsadat 2015 | [9] |
| Freynhagen 2005 | [11] |
| Gao 2010 | [10] |
| Gao 2015 | [12] |
| Goldstein 2005 | [13] |
| Graff 2000 | [14] |
| Guan 2011 | [15] |
| Hui 2011 | [16] |
| Irving 2011 | [17] |
| Jang 2017 | [18] |
| Jiang 2019 | [19] |
| Kieburtz 1998 | [21] |
| Liu 2014 | [20] |
| Liu 2015 | [22] |
| Markman 2018 | [23] |
| Mathieson 2017 | [24] |
| Moon 2010 | [25] |
| Mu 2017 | [26] |
| NCT 00394901 | [27] |
| Raskin 2005 | [28] |
| Rauck 2012 | [29] |
| Rosenstock 2004 | [30] |
| Rowbotham 1998 | [31] |
| Sabatowski 2004 | [32] |
| Sang 2013 | [33] |
| Satoh 2010 | [34] |
| Serpell 2002 | [35] |
| Shlay 1998 | [39] |
| Simpson 2001 | [36] |
| Simpson 2008 | [40] |
| Simpson 2010 | [37] |
| Simpson 2014 | [38] |
| Simpson 2017 | [41] |
| Tolle 2008 | [42] |
| van Seventer 2006 | [43] |
| van Seventer 2010 | [44] |
| Wallace 2010 | [45] |
| Webster 2010a | [46] |
| Webster 2010b | [47] |
| Wernicke 2006 | [48] |
| Yasuda 2011 | [49] |
| Zhang 2013 | [50] |

[1] Allen R, Sharma U, Barlas S. Clinical experience with desvenlafaxine in treatment of pain associated with diabetic peripheral neuropathy. Journal of Pain Research 2014;7:339-351.

[2] Arezzo JC, Rosenstock J, LaMoreaux L, Pauer L. Efficacy and safety of pregabalin 600 mg/d for treating painful diabetic peripheral neuropathy: A double-blind placebo-controlled trial. BMC Neurol 2008;8.

[23] Markman J, Resnick M, Greenberg S, Katz N, Yang R, Scavone J, Whalen E, Gregorian G, Parsons B, Knapp L. Efficacy of pregabalin in post-traumatic peripheral neuropathic pain: a randomized, double-blind, placebo-controlled phase 3 trial. Journal of neurology, 2018.

[47] Webster LR, Tark M, Rauck R, Tobias JK, Vanhove GF. Effect of duration of postherpetic neuralgia on efficacy analyses in a multicenter, randomized, controlled study of NGX-4010, an 8% capsaicin patch evaluated for the treatment of postherpetic neuralgia. BMC Neurol 2010;10.

[48] Wernicke JF, Pritchett YL, D'Souza DN, Waninger A, Tran P, Iyengar S, Raskin J. A randomized controlled trial of duloxetine in diabetic peripheral neuropathic pain. Neurology 2006;67(8):1411-1420.

[49] Yasuda H, Hotta N, Nakao K, Kasuga M, Kashiwagi A, Kawamori R. Superiority of duloxetine to placebo in improving painful diabetic neuropathic pain: results of a randomized controlled trial in Japan. Journal of diabetes investigation, 2011. pp. 132‐139.

[50] Zhang L, Rainka M, Freeman R, Harden RN, Bell CF, Chen C, Graff O, Harding K, Hunter S, Kavanagh S, Laurijssens B, Schwartzbach C, Warren S, McClung C. A Randomized, Double-Blind, Placebo-Controlled Trial to Assess the Efficacy and Safety of Gabapentin Enacarbil in Subjects with Neuropathic Pain Associated with Postherpetic Neuralgia (PXN110748). JPain 2013;14(6):590-603.

### List of excluded studies

| Author | DOI/NCT reference | Reason |
| --- | --- | --- |
| Achar 2010 | 10.4103/0378-6323.58686 | No usable data |
| Achar 2012 | NA | No usable data |
| Achar 2013 | NA | No usable data |
| Adams 2016 | 10.1177/1740774516631530 | Intervention |
| Al-Hihi 2017 | 10.7326/ACPJC-2017-167-2-004 | Study design |
| Aldrete 2000 | NA | Study design |
| Alpizar 2012 | https://clinicaltrials.gov/ct2/show/NCT01364298 | Duplicate |
| Arai 2010 | 10.1007/s00540-010-0913-6 | Length of follow-up |
| Asilian 2002 | NA | Length of follow-up |
| Atalay 2013 | 10.1007/s40261-013-0080-2 | Length of follow-up |
| Athanasakis 2013 | 10.1186/1471-2377-13-56 | Study design |
| Athanasakis 2013 | 10.1186/1471-2377-13-56. | Study design |
| Atli 2005 | 10.1111/j.1526-4637.2005.05035.x | Intervention |
| Avan 2018 | 10.4103/jrms.JRMS_1068_17 | Length of follow-up |
| Babar Melik 2015 | NA | Population |
| Backonja 1999 | 10.1111/j.1528-1157.1999.tb00934.x | No usable data |
| Backonja 2003 | NA | Study design |
| Backonja 2008 | NA | Duplicate |
| Backonja 2010 | 10.1111/j.1526-4637.2009.00793.x | Duplicate |
| Backonja 2010 | 10.1016/S1754-3207(10)70529-0 | Study design |
| Backonja 2011 | 10.1111/j.1526-4637.2011.01139.x. | Length of follow-up |
| Backonja 2012 | 10.1111/j.1526-4637.2012.01331.x | Study design |
| Backryd 2015 | 10.1111/ner.12293 | Intervention |
| Banerjee 2017 | 10.1007/s11606-017-4028-8 | Study design |
| Bansal 2009 | 10.1111/j.1464-5491.2009.02806.x. | Length of follow-up |
| Barbarisi 2010 | 10.1097/AJP.0b013e3181dda1ac | Comparator |
| Barnhoorn 2015 | 10.1136/bmjopen-2015-008283 | Intervention |
| Baron 2009 | 10.1185/03007990903048078 | Study design |
| Baron 2009a | NA | Length of follow-up |
| Baron 2009b | 10.2165/00044011-200929040-00002 | Length of follow-up |
| Baron 2009c | NA | Length of follow-up |
| Baron 2010 | 10.1002/central/CN-00690009 | Length of follow-up |
| Baron 2010 | 10.1016/j.pain.2010.04.013 | Length of follow-up |
| Baron 2015 | 10.1111/papr.12200 | Comparator |
| Barrett 2008 | 10.1016/j.jpain.2008.01.205 | Length of follow-up |
| Barton 2011 | 10.1007/s00520-010-0911-0 | Intervention |
| Battaglini 2018 | 10.1016/j.cct.2018.04.011 | Study ongoing |
| Battla 1981 | NA | Study design |
| Belfer 2017 | 10.1097/PR9.0000000000000596 | Study design |
| Bernstein 1989 | 10.1016/S0190-9622(89)70171-7 | Length of follow-up |
| Bhaskar 2012 | NA | Study design |
| Bhat 2014 | NA | Length of follow-up |
| Biesbroeck 1995 | NA | No usable data |
| Biyik 2009 | NA | Length of follow-up |
| Biyik 2012 | 10.1007/s11255-012-0193-1. | Length of follow-up |
| Block 2009 | NA | Study design |
| Blonna 2004 | NA | Study design |
| Bouhassira 2014 | 10.1016/j.pain.2014.08.020 | Study design |
| Boulton 2011 | 10.1007/s11892-011-0199-6 | Study design |
| Bowsher 1994 | NA | No usable data |
| Bowsher 1997 | NA | POPULATION |
| Boyle 2012 | NA | Length of follow-up |
| Brasch-Andersen 2010 | NA | Study design |
| Brasch-Andersen 2011 | 10.1007/s00228-011-1056-x | Study design |
| Brogly 2008 | 10.1213/ane.0b013e318185cf73 | Population |
| Bruce 2014 | NA | Study design |
| Calkins 2014 | 10.1016/j.jpain.2014.01.298 | Duplicate |
| Calkins 2014 | 10.1016/j.jpain.2014.01.299 | Duplicate |
| Calkins 2014 | 10.1016/j.pmrj.2014.08.891 | Duplicate |
| Calkins 2016 | 10.1093/pm/pnv072 | Duplicate |
| Campbell 1966 | 10.1136/jnnp.29.3.265 | Length of follow-up |
| Campbell 2011 | NA | Study design |
| Canovas 2009 | NA | Study design |
| Caraceni 2004 | 10.1200/JCO.2004.08.141 | Length of follow-up |
| Carasso 1979 | NA | Comparator |
| Cartagena 2005 | NA | Study design |
| Chan 2013 | NA | No usable data |
| Chappell 2005 | NA | Endpoints |
| Chaudhry 2017 | 10.2217/pgs-2016-0185 | Study design |
| Choudhary 2018 | 10.4103/idoj.IDOJ_377_16 | POPULATION |
| Ciaramella 2000 | NA | Study design |
| Cohen 2015 | 10.1097/ALN.0000000000000409 | Intervention |
| Cvijanovic 2017 | 10.2298/VSP151209261C | Study design |
| Dailey 1992 | 10.2337/diacare.15.2.159 | Intervention |
| Dallocchio 2001 | NA | Duplicate |
| Daniel 2013 | NA | Length of follow-up |
| De Heer 2013 | 10.1186/1471-244X-13-147 | No usable data |
| De Heer 2018 | 10.3389/fpsyt.2018.00118 | Population |
| de Jaeger 2018 | NA | No usable data |
| Demitrack 2003 | NA | No usable data |
| Dinat 2010 | 10.1016/S1754-3207(10)70491-0 | Length of follow-up |
| Dinat 2015 | doi:10.1371/journal. pone.0126297 | Length of follow-up |
| Ding 2014 | 10.3969/j.issn.1000-8179.20141212 | COMPARATOR |
| Dini 1993 | 10.1016/0304-3959(93)90213-9 | Intervention |
| Dolgun 2014 | 10.1179/1743132814Y.0000000404 | Study design |
| Donofrio 1991 | 10.1001/archinte.1991.00400110079017 | Intervention |
| Dou 2014 | 10.1111/ajco.12311 | Length of follow-up |
| Drake 1990 | NA | No usable data |
| Dunteman 2015 | NA | Duplicate |
| Durand 2012 | NA | Population |
| Dworkin 2007 | https://doi.org/10.1017/S1748232107000080 | Study design |
| Dworkin 2009 | 10.1016/j.pain.2008.12.022 | POPULATION |
| Ebell 2017 | NA | No usable data |
| Edwards 1998 | NA | No usable data |
| Eerdekens 2016 | NA | Intervention |
| Ehrnrooth 2001 | NA | Population |
| Elchami 2011 | https://doi.org/10.1016/S1754-3207(11)70950-6 | Intervention |
| Elchami 2014 | NA | Study design |
| Ellison 1997 | 10.1200/JCO.1997.15.8.2974 | Intervention |
| Ermis 2010 | 10.1016/j.jdiacomp.2008.12.001 | Population |
| Farrar 2014 | 10.1016/j.pain.2014.05.009 | Study design |
| Farshchian 2018 | NA | Length of follow-up |
| Frank 2008 | 10.2217/14796708.3.6.631 | Study design |
| Freeman 2015 | 10.1111/pme.12791 | Study design |
| Freynhagen 2005 | 10.1016/j.pain.2005.02.032 | Population |
| Freynhagen 2006 | 10.1007/s00482-005-0449-0 | Study design |
| Gabrani 2016 | NA | POPULATION |
| Galindo 2012 | https://doi.org/10.1016/j.jval.2011.08.1569 | Comparator |
| Gatti 2008 | 10.1159/000186502 | Study design |
| Gelijkens 2014 | NA | Length of follow-up |
| Gerson 1977 | NA | Study design |
| Gewandter 2014 | NA | Intervention |
| Ghai 2011 | 10.4103/1658-354X.84097 | Population |
| Gilron 2005 | 10.1056/NEJMoa042580 | Length of follow-up |
| Gilron 2009 | doi:10.1016/s0140-6736(09)61081-3 | Length of follow-up |
| Gilron 2011 | doi:10.1097/ajp.0b013e3181fe13f6 | Length of follow-up |
| Gilron 2013 | NA | Study design |
| Gilron 2015 | 10.1097/j.pain.0000000000000149 | Length of follow-up |
| Gobel 1995 | NA | Study design |
| Gobel 1997 | NA | Study design |
| Goldman 2010 | 10.1016/j.pain.2010.01.016 | Population |
| Goldstein 2003 | NA | Duplicate |
| Gomez Perez 1985 | NA | Length of follow-up |
| Gomez Perez 1996 | NA | Length of follow-up |
| Gomez Perez 2004 | NA | Length of follow-up |
| Gong 2008 | NA | Length of follow-up |
| Gonzalez Duarte 2016 | 10.1097/AJP.0000000000000339 | Length of follow-up |
| Gordh 2008 | 10.1016/j.pain.2007.12.011 | Length of follow-up |
| Gorson 1999 | NA | Length of follow-up |
| Gray 2009 | NA | Length of follow-up |
| Gray 2011 | 10.1016/j.pain.2011.01.055 | Length of follow-up |
| Gribble 2009 | NA | Length of follow-up |
| Gribble 2010 | NA | Length of follow-up |
| Griebeler 2014 | 10.7326/M14-0511 | Study design |
| Grosskopf 2006 | 10.1111/j.1600-0404.2005.00559.x | Intervention |
| Gu 2012 | NA | No usable data |
| Guan 2011 | 10.1016/j.clinthera.2011.02.007 | No usable data |
| Haanpaa 2014 | NA | Study design |
| Hahn 2004 | 10.1007/s00415-004-0529-6 | Length of follow-up |
| Hambardzumyan 2017 | 10.1016/j.jns.2017.08.2749 | Intervention |
| Hammack 2002 | NA | Length of follow-up |
| Hanks 1981 | 10.1016/0304-3959(81)90319-5 | Study design |
| Harden 2013 | 10.1111/pme.12227 | Length of follow-up |
| Hardy 2007 | 10.2337/dc06-0947 | Study design |
| Harke 2001 | 10.1213/00000539-200102000-00039 | Intervention |
| Harrison 2013 | 10.1111/pme.12084 | Length of follow-up |
| Hasani 2009 | NA | Length of follow-up |
| Hassan 2013 | NA | Length of follow-up |
| Hazaert 2012 | 10.1016/j.ymgme.2011.11.075 | No usable data |
| Heras 2016 | NA | Length of follow-up |
| Hewitt 2009 | DOI: https://doi.org/10.1016/j.jpain.2009.01.171 | Length of follow-up |
| Hewitt 2011 | 10.1016/j.pain.2010.10.050 | Length of follow-up |
| Hirayama 2015 | NA | Length of follow-up |
| Ho 2008 | 10.1097/AJP.0b013e318156db26 | Intervention |
| Ho 2009 | 10.1016/j.pain.2008.07.013 | Length of follow-up |
| Hoffman 2009 | 10.1016/j.pain.2009.09.017 | No usable data |
| Holbech 2015 | 10.1097/j.pain.0000000000000143 | Length of follow-up |
| Hoseinzade 2008 | NA | No usable data |
| Hota 2009 | NA | Length of follow-up |
| Huffman 2015 | 10.1097/AJP.0000000000000198 | Length of follow-up |
| Hutmacher 2016 | 10.1002/jcph.567 | Length of follow-up |
| Iacobellis 2000 | NA | Population |
| Imani 2009 | NA | Length of follow-up |
| Irving 2012 | 10.2147/JPR.S32562 | Study design |
| Irving 2009 | 10.1097/AJP.0b013e3181934276 | Length of follow-up |
| Irving 2010 | NA | Intervention |
| Irving 2012 | NA | Duplicate |
| Jann 2007 | NA | Study design |
| Jean 2005 | NA | No usable data |
| Jenkins 2010 | 10.1016/S1754-3207(10)70314-X | Length of follow-up |
| Jenkins 2012 | 10.2147/JPR.S34098 | Length of follow-up |
| Jensen 2009 | 10.1097/AJP.0b013e318192bf87. | Length of follow-up |
| Jensen 2012 | 10.1016/j.jpain.2012.01.290 | Study design |
| Jensen 2012 | 10.1111/j.1526-4637.2012.01427.x | Study design |
| Jensen 2012 | 10.1097/AJP.0b013e31823f9e64 | Length of follow-up |
| Jensen 2013 | 10.1097/AJP.0b013e31827b32ab. | Study design |
| Jensen 2014 | 10.1002/j.1532-2149.2014.00479.x | Length of follow-up |
| Ji 2015 | NA | Study design |
| Jia 2006 | NA | Length of follow-up |
| Jiang 2011 | 10.1097/JCP.0b013e31820f4f57 | Length of follow-up |
| Jose 2007 | 10.1111/j.1464-5491.2007.02093.x | Length of follow-up |
| Kalita 2014 | 10.1016/j.jns.2014.05.002 | Population |
| Kalliomaki 2013 | 10.1016/j.sjpain.2012.10.003 | Intervention |
| Kalso- eija 1995 | NA | Length of follow-up |
| Kanodia 2011 | 10.5214/ans.0972.7531.1118405 | POPULATION |
| Kanodia 2012 | 10.4103/0019-5154.100476 | POPULATION |
| Kantito 2014 | NA | Population |
| Kantor 2012 | NA | No usable data |
| Kardanpour 2018 | 10.22122/jims.v35i462.9005 | Population |
| Karmakar 2014 | 10.1186/1743-0003-11-125 | Length of follow-up |
| Kartapol'tseva 2011 | NA | Study design |
| Kaur 2011 | 10.2337/dc10-1793 | Length of follow-up |
| Kautio 2009 | NA | Population |
| Kerckhove 2018 | 10.1002/ejp.1221 | Intervention |
| Keskinbora 2006 | NA | Length of follow-up |
| Keskinbora 2007 | NA | Length of follow-up |
| Khan 2014 | 10.1117/12.2036376? | Comparator |
| Khoromi 2007 | 10.1016/j.pain.2006.10.029 | Comparator |
| Kiani 2015a | NA | Intervention |
| Kiani 2015b | NA | Intervention |
| Kieburtz 1998 | 10.1212/WNL.51.6.1682 | No usable data |
| Kishore-Kumar 1990 | 10.1002/central/CN-00066244 | Length of follow-up |
| Knight 1994 | 10.1111/j.1365-4362.1994.tb04955.x | Study design |
| Ko 2010 | 10.1111/j.1464-5491.2010.03054.x | Length of follow-up |
| Krcevski 2010 | 10.1002/central/CN-00760796 | Length of follow-up |
| Kulkantrakorn 2012 | 10.1111/papr.12013 | Intervention |
| Kulkantrakorn 2018 | 10.1016/j.jocn.2018.11.036 | Intervention |
| Kunz 2000 | 10.1016/S0924-977X(00)80539-8 | Length of follow-up |
| Kunz 2002 | NA | Duplicate |
| Kvinesdal 1984 | 10.1001/jama.1984.03340370059031 | Length of follow-up |
| Kwasucki 2002 | NA | Length of follow-up |
| Langohr 1982 | 10.1159/000115497 | Length of follow-up |
| Lee 2012 | 10.3928/01477447-20120525-21. | Study design |
| Lee 2016 | 10.1111/dth.12331 | POPULATION |
| Lemos 2008 | 10.1097/AJP.0b013e318158011a | Length of follow-up |
| Lemos 2010 | 10.2147/JPR.S13154 | Length of follow-up |
| Lemos 2011 | 10.2147/JPR.S20555 | Study design |
| Lesser 2004 | 10.1212/01.WNL.0000145767.36287.A1 | Length of follow-up |
| Liang 2015 | 10.1002/central/CN-01048935 | Population |
| Liu 2015 | 10.1002/central/CN-01434810/full | Duplicate |
| Llobera 2013 | 10.3109/13814788.2012.759936 | POPULATION |
| Lo 2014 | 10.4021/jocmr879w | Study design |
| Low 1995 | 10.1016/0304-3959(94)00261-C | Intervention |
| Lynch 2003 | NA | Intervention |
| Lynch 2005 | NA | Intervention |
| Mailis 1997 | 10.1016/S0304-3959(96)03300-3 | Intervention |
| Malik 2015 | 10.5812/aapm.28110 | Length of follow-up |
| Mankowski 2017 | 10.1186/s12883-017-0836-z. | Study design |
| Marcus 2018 | 10.1155/2018/2140420 | Study design |
| Marinelli 2009 | NA | Intervention |
| Martini 2012 | doi: 10.2147/JPR.S30406 | Study design |
| Mathieson 2016 | https://doi.org/10.1186/s13063-016-1174-y | No usable data |
| Mathis 2012 | 10.1331/JAPhA.2012.12510 | No usable data |
| Matsuoka 2017 | NA | Length of follow-up |
| Matsuoka 2018 | NA | Population |
| Max 1987 | NA | Length of follow-up |
| Max 1988 | NA | Length of follow-up |
| Max 1991 | 10.1016/0304-3959(91)90157-S | Length of follow-up |
| Max 1992 | 10.1056/NEJM199205073261904 | Length of follow-up |
| McCleane 2000 | 10.1046/j.1365-2125.2000.00200.x | Length of follow-up |
| Medina-Santillan 2004 | NA | Duplicate |
| Medvedeva 2008 | NA | Intervention |
| Mendel 1986 | 10.1001/jama.1986.03370050079025 | Length of follow-up |
| Merante 2014 | 10.2337/db14-389-664 | Length of follow-up |
| Merante 2017 | NA | Duplicate |
| Merante 2017 | 10.1093/pm/pnw342 | Length of follow-up |
| Mercadante 2002 | NA | Length of follow-up |
| Meurant 2006 | 0.1186/cc4786 | Population |
| Milenkovic 2009 | NA | Study design |
| Minotti 1998 | NA | Length of follow-up |
| Mishra 2012 | NA | Length of follow-up |
| Mishra 2013 | 10.7860/JCDR/2013/5707.3239 | Length of follow-up |
| Misiego 2000 | NA | Study design |
| Miyazaki 2016 i | NA | Population |
| Moch 2010 | 10.1111/j.1742-7843.2010.00600.x | No usable data |
| Molina 2007 | NA | Population |
| Moon 2017 | NA | Intervention |
| Moore 2018 | 10.1001/jama.2017.21547 | Study design |
| Morello 1999 | 10.1001/archinte.159.16.1931 | Length of follow-up |
| Morgenlander 1990 | 10.1002/ana.410280222 | Study design |
| Musharraf 2017 | NA | Intervention |
| Najafi 2018 | 10.1016/j.burns.2018.10.011 | Length of follow-up |
| Nasare 2015 | 10.13040/IJPSR.0975-8232.6(4).1568-78 | Length of follow-up |
| Nazarbaghi 2017 | 10.19082/5617 | Length of follow-up |
| Nct 00085761 | Https://clinicaltrials.gov/show/nct00085761 | No usable data |
| NCT 00159705 | Https://clinicaltrials.gov/show/nct00159705 | No usable data |
| NCT 00904202 | Www.clinicaltrials.gov/show/nct00904202 | Length of follow-up |
| Nct 1999 | Https://clinicaltrials.gov/show/nct00004390 | No usable data |
| Nct 2003 | Https://clinicaltrials.gov/show/nct00061776 | No usable data |
| Nct 2005 | Https://clinicaltrials.gov/show/nct00159666 | No usable data |
| Nct 2005 | Https://clinicaltrials.gov/show/nct00143442 | No usable data |
| Nct 2005 | Https://clinicaltrials.gov/show/nct00159640 | No usable data |
| Nct 2006 | Https://clinicaltrials.gov/show/nct00334685 | No usable data |
| Nct 2007 | Https://clinicaltrials.gov/show/nct00475904 | Length of follow-up |
| Nct 2008 | Https://clinicaltrials.gov/show/nct00599638 | No usable data |
| Nct 2008 | Https://clinicaltrials.gov/show/nct00619476 | Duplicate |
| Nct 2008 | Https://clinicaltrials.gov/show/nct00636636 | Duplicate |
| Nct 2010 | Https://clinicaltrials.gov/show/nct01058642 | Length of follow-up |
| Nct 2011 | Https://clinicaltrials.gov/show/nct01455428 | Duplicate |
| Nct 2012 | Https://clinicaltrials.gov/show/nct01713426 | No usable data |
| Nct 2015 | Https://clinicaltrials.gov/show/nct02868801 | No usable data |
| Nct 2017 | Https://clinicaltrials.gov/show/nct03186443 | No usable data |
| NCT00027963 | Https://clinicaltrials.gov/show/nct00027963 | No usable data |
| NCT00143156 | Https://clinicaltrials.gov/show/nct00143156 | No usable data |
| NCT00156078 | Https://clinicaltrials.gov/show/nct00156078 | No usable data |
| NCT00162968 | Https://clinicaltrials.gov/show/nct00162968 | Length of follow-up |
| NCT00189072 | Https://clinicaltrials.gov/show/nct00189072 | Length of follow-up |
| NCT00266643 | Https://clinicaltrials.gov/show/nct00266643 | No usable data |
| NCT00283842 | Https://clinicaltrials.gov/show/nct00283842 | Duplicate |
| NCT00291148 | https://clinicaltrials.gov/ct2/show/NCT00291148 | No usable data |
| NCT00301223 | Https://clinicaltrials.gov/show/nct00301223 | No usable data |
| NCT00322621 | Https://clinicaltrials.gov/show/nct00322621 | Study design |
| NCT00380874 | Https://clinicaltrials.gov/show/nct00380874 | Population |
| NCT00480181 | https://clinicaltrials.gov/ct2/show/NCT00480181 | Intervention |
| NCT00507936 | Https://clinicaltrials.gov/show/nct00507936 | No usable data |
| NCT00516503 | Https://clinicaltrials.gov/show/nct00516503 | Intervention |
| NCT00553475 | Https://clinicaltrials.gov/show/nct00553475 | Duplicate |
| NCT00570310 | Https://clinicaltrials.gov/show/nct00570310 | Length of follow-up |
| NCT00619983 | Https://clinicaltrials.gov/show/nct00619983 | Length of follow-up |
| NCT00634543 | Https://clinicaltrials.gov/show/nct00634543 | Length of follow-up |
| NCT00654940 | Https://clinicaltrials.gov/show/nct00654940 | Length of follow-up |
| NCT00740571 | Https://clinicaltrials.gov/show/nct00740571 | Study design |
| NCT00772291 | NCT00772291 | No usable data |
| NCT00785577 | Https://clinicaltrials.gov/show/nct00785577 | Length of follow-up |
| NCT00837941 | Https://clinicaltrials.gov/show/nct00837941 | No usable data |
| NCT00844194 | Https://clinicaltrials.gov/show/nct00844194 | Study design |
| NCT00852436 | Https://clinicaltrials.gov/show/nct00852436 | No usable data |
| NCT00908375 | Https://clinicaltrials.gov/show/nct00908375 | Length of follow-up |
| NCT00944697 | Https://clinicaltrials.gov/show/nct00944697 | Intervention |
| NCT00967707 | Https://clinicaltrials.gov/show/nct00967707 | No usable data |
| NCT00993070 | Https://clinicaltrials.gov/show/nct00993070 | Intervention |
| NCT01047488 | Https://clinicaltrials.gov/show/nct01047488 | Length of follow-up |
| NCT01057693 | Https://clinicaltrials.gov/show/nct01057693 | Study design |
| NCT01067144 | Https://clinicaltrials.gov/show/nct01067144 | Population |
| NCT01089556 | Https://clinicaltrials.gov/show/nct01089556 | Duplicate |
| NCT01125215 | Https://clinicaltrials.gov/show/nct01125215 | Intervention |
| NCT01127100 | Https://clinicaltrials.gov/show/nct01127100 | No usable data |
| NCT01166048 | Https://clinicaltrials.gov/show/nct01166048 | No usable data |
| NCT01177514 | NCT01177514 | No usable data |
| NCT01210079 | Https://clinicaltrials.gov/show/nct01210079 | Length of follow-up |
| NCT01225068 | Clinicaltrials.gov/ct2/show/record/nct01225068 | Length of follow-up |
| NCT01263132 | Https://clinicaltrials.gov/show/nct01263132 | Length of follow-up |
| NCT01288937 | Https://clinicaltrials.gov/show/nct01288937 | No usable data |
| NCT01314222 | Https://clinicaltrials.gov/show/nct01314222 | No usable data |
| NCT01332149 | Https://clinicaltrials.gov/show/nct01332149 | Duplicate |
| NCT01352741 | Https://clinicaltrials.gov/show/nct01352741 | Intervention |
| NCT01416116 | Https://clinicaltrials.gov/show/nct01416116 | Intervention |
| NCT01455415 | Https://clinicaltrials.gov/show/nct01455415 | Length of follow-up |
| NCT01474772 | Https://clinicaltrials.gov/show/nct01474772 | Length of follow-up |
| NCT01485094 | Https://clinicaltrials.gov/show/nct01485094 | Length of follow-up |
| NCT01504412 | Https://clinicaltrials.gov/show/nct01504412 | Length of follow-up |
| NCT01533428 | Https://clinicaltrials.gov/show/nct01533428 | Duplicate |
| NCT01556152 | Https://clinicaltrials.gov/show/nct01556152 | Length of follow-up |
| NCT01579279 | Https://clinicaltrials.gov/show/nct01579279 | No usable data |
| NCT01588314 | Https://clinicaltrials.gov/show/nct01588314 | No usable data |
| NCT01611155 | Https://clinicaltrials.gov/show/nct01611155 | Population |
| NCT01637077 | Https://clinicaltrials.gov/show/nct01637077 | Population |
| NCT01770964 | Https://clinicaltrials.gov/show/nct01770964 | Length of follow-up |
| NCT01821430 | Https://clinicaltrials.gov/show/nct01821430 | No usable data |
| NCT01838044 | Https://clinicaltrials.gov/show/nct01838044 | Intervention |
| NCT01863810 | Https://clinicaltrials.gov/show/nct01863810 | No usable data |
| NCT01869569 | Https://clinicaltrials.gov/show/nct01869569 | No usable data |
| NCT01928381 | Https://clinicaltrials.gov/show/nct01928381 | Length of follow-up |
| NCT01939366 | Https://clinicaltrials.gov/show/nct01939366 | Length of follow-up |
| NCT02074267 | Https://clinicaltrials.gov/show/nct02074267 | Length of follow-up |
| NCT02215252 | Https://clinicaltrials.gov/show/nct02215252 | Length of follow-up |
| NCT02394951 | Https://clinicaltrials.gov/show/nct02394951 | Length of follow-up |
| NCT02417935 | Https://clinicaltrials.gov/show/nct02417935 | Duplicate |
| NCT02607254 | Https://clinicaltrials.gov/show/nct02607254 | Population |
| NCT02673866 | Https://clinicaltrials.gov/show/nct02673866 | Length of follow-up |
| NCT02822625 | Https://clinicaltrials.gov/show/nct02822625 | Intervention |
| NCT02869867 | Https://clinicaltrials.gov/show/nct02869867 | Intervention |
| NCT02927951 | Https://clinicaltrials.gov/show/nct02927951 | Length of follow-up |
| NCT02985216 | Https://clinicaltrials.gov/show/nct02985216 | No usable data |
| NCT03113448 | Https://clinicaltrials.gov/show/nct03113448 | Intervention |
| NCT03202979 | Https://clinicaltrials.gov/show/nct03202979 | No usable data |
| NCT03324035 | Https://clinicaltrials.gov/show/nct03324035 | No usable data |
| NCT03348735 | Https://clinicaltrials.gov/show/nct03348735 | No usable data |
| Nicol 1969 | 10.1111/j.1526-4610.1969.hed0901054.x | Intervention |
| Nocivin 2003 | 10.1046/j.1468-3083.17.s3.8.x | POPULATION |
| Otsuki 2016 | 10.1007/s40261-016-0464-1. | Study design |
| Otto 2009 | 10.1016/j.pain.2008.04.012 | Length of follow-up |
| Paech 2007 | 10.1213/01.ane.0000286227.13306.d7 | Population |
| Paice 2000 | 10.1016/S0885-3924(99)00139-6 | Length of follow-up |
| Paladini 1987 | NA | Study design |
| Palomba 2009 | 10.1016/S1090-3801(09)60578-4 | Length of follow-up |
| Pan 2008 | NA | Intervention |
| Pandey 2005 | 10.1213/01.ANE.0000152186.89020.36 | Length of follow-up |
| Papaioannou 2018 | NA | Comparator |
| Park 2009 | 10.1111/j.1529-8027.2009.00236.x | Length of follow-up |
| Parsons 2015 | 10.1002/ana.24498 | Duplicate |
| Parsons 2018 | 10.2147/JPR.S157856 | Study design |
| Pasnoor 2018 | NA | Duplicate |
| Patarica-Huber 2011 | NA | Length of follow-up |
| Patel 2012 | 10.1331/JAPhA.2012.12510 | Study design |
| Pelloso 2005 | 10.11606/T.5.2005.tde-06022007-113055 | Population |
| Penide 2012 | NA | Study design |
| Perez 2000 | 10.1016/S0002-9343(00)00398-3 | Study design |
| Pi 2018 | 10.3760/cma.j.issn.0376-2491.2018.10.004. | Intervention |
| Portilla 2013 | 10.1097/BCR.0b013e3182700675 | Intervention |
| Pritchett 2007 | 10.1111/j.1526-4637.2007.00305.x | Study design |
| Psurek 2011 | NA | Length of follow-up |
| Raja 2002 | 10.1212/WNL.59.7.1015 | No usable data |
| Rajanandh 2014 | 10.1016/j.pharep.2013.08.003 | Intervention |
| Rao 2007 | NA | Length of follow-up |
| Raptis 2013 | NA | Length of follow-up |
| Raskin 2006a | 10.1089/jpm.2006.9.29 | Study design |
| Raskin 2006b | 10.1111/j.1526-4637.2006.00207.x | Intervention |
| Raskin 2014 | 10.1097/AJP.0b013e31829ea1a1 | Length of follow-up |
| Raskin 2016 | 10.1097/AJP.0000000000000254 | Length of follow-up |
| Rauck 2010 | NA | Duplicate |
| Rauck 2013 | 10.1097/ALN.0b013e3182a10fbf | Intervention |
| Rawn 2000 | NA | Length of follow-up |
| Razazian 2014 | NA | Length of follow-up |
| Rehm 2010 | 10.1185/03007995.2010.483675 | Length of follow-up |
| Ren 2016 | NA | Length of follow-up |
| Reyad 2019 | 10.1016/j.jpainsymman.2018.10.496 | Population |
| Rice 2001 | NA | Length of follow-up |
| Richter 2005 | 10.1016/j.jpain.2004.12.007 | Length of follow-up |
| Robbins 1998 | 10.1213/00000539-199803000-00027 | Intervention |
| Robertson 2016 | 10.1093/pm/pnw052 | Study design |
| Robertson 2018 | 10.1186/s13063-017-2400-y | Study design |
| Rockliff 1966 | 10.1001/archneur.1966.00470140019003 | Length of follow-up |
| Romanò 2009 | 10.1007/s10195-009-0077-z | Length of follow-up |
| Rossignol 2019 | NA | Intervention |
| Rovetta 1999 | NA | Study design |
| Rowbotham 2004 | 10.1016/j.pain.2004.05.010 | Length of follow-up |
| Rowbotham 2005 | 10.1016/j.jpain.2005.07.001 | Length of follow-up |
| Rowbotham 2005 | NA | Duplicate |
| Rowbotham 2009 | 10.1111/j.1533-2500.2009.00266.x | Duplicate |
| Roy 2013 | NA | Population |
| Rull 1969 | NA | Intervention |
| Rullán 2017 | 10.1186/s13063-016-1729-y | Study design |
| Rustagi 2014 | 10.1007/s12663-013-0513-8. | Length of follow-up |
| Sabatowski 2004 | 10.1111/j.1468-1331.2004.00919.x | Duplicate |
| Sakai 2015 | 10.1007/s00586-015-3812-6 | Length of follow-up |
| Sandercock 2009 | 10.2337/dc08-1450 | Length of follow-up |
| Sandercock 2012 | 10.1016/j.diabres.2012.03.010 | Length of follow-up |
| Sardar 2018 | NA | Intervention |
| Satoh 2011 | 10.1111/j.2040-1124.2011.00122.x | Study design |
| Satomi 2018 | NA | Population |
| Saxena 2016 | NA | Intervention |
| Scheffler 1991 | 10.7547/87507315-81-6-288 | Intervention |
| Schliessbach 2018 | https://doi.org/10.1371/journal.pone.0195776 | Length of follow-up |
| Schug 2017 | NA | Study design |
| Schukro 2016 | 10.1097/ALN.0000000000000902 | Length of follow-up |
| Seidl 1999 | NA | Study design |
| Selvarajah 2018 | 10.1186/s13063-018-2959-y | No usable data |
| Serpell 2010 | NA | Length of follow-up |
| Serpell 2010 | 10.1016/S1754-3207(10)70313-8 | No usable data |
| Shabbir 2011 | NA | Length of follow-up |
| Sharma 2005 | NA | Study design |
| Sharma 2006 | NA | Duplicate |
| Shim 2010 | 10.1200/jco.2010.28.15_suppl.tps315 | Comparator |
| Shinde 2015 d | NA | Population |
| Shurman 2015 | NA | No usable data |
| Silver 2007 | NA | Population |
| Silver 2007 | 10.1016/j.jpainsymman.2006.12.015 | Intervention |
| Sima 2011 | NA | No usable data |
| Simpson 2009 | NA | No usable data |
| Simpson 2017 | 0.1016/j.jpain.2016.09.008 | Duplicate |
| Sindrup 1989 | NA | Length of follow-up |
| Sindrup 1990 | NA | Length of follow-up |
| Sindrup 1992a | NA | Length of follow-up |
| Sindrup 1992b | 10.1038/clpt.1992.183 | Length of follow-up |
| Sindrup 1992c | 10.1007/BF02333018 | Length of follow-up |
| Sindrup 2003 | https://doi.org/10.1212/01.WNL.0000058749.49264.BD | Length of follow-up |
| Sindrup 2017 | NA | Study design |
| Singh 2013 | 10.1111/pme.12001 | Intervention |
| Skvarc 2009 | NA | POPULATION |
| Skvarc 2010 | 10.1007/s00508-010-1345-x | POPULATION |
| Smith 2013 | NA | Length of follow-up |
| Smugar 2011 | https://doi.org/10.1016/S1754-3207(11)70186-9 | Length of follow-up |
| Snijder 2015 | NA | Duplicate |
| Solak 2007 | 10.1016/j.ejcts.2007.03.022 | Study design |
| Solaro 1998 | 10.1212/wnl.51.2.609 | COMPARATOR |
| Stacey 2008 | 10.1016/j.jpain.2008.05.014 | Length of follow-up |
| Stacey 2008 | 10.1111/j.1526-4637.2008.00423.x | Study design |
| Steigerwald 2013 | NA | ComPARATOR |
| Stocker 2015 | 10.1007/s00125-015-3687-4 | Duplicate |
| Strojek 2004 | https://doi.org/10.1016/j.jpain.2004.02.204 | Duplicate |
| Stump 2009 | NA | Study design |
| Sumracki 2012 | 10.1371/journal.pone.003852 | Intervention |
| Sun 2011 | NA | Length of follow-up |
| Sweeney 2012 | NA | Study design |
| Sweeney 2014 | NA | Study design |
| Talaei 2009 | 10.1080/09637480802406153 | Comparator |
| Tamez-Perez 1998 | NA | Study design |
| Tanaka 2016 | 10.1016/j.jdermsci.2016.08.111 | POPULATION |
| Tandan 1992 | 10.2337/diacare.15.1.8 | Intervention |
| Tanenberg 2011b | i10.4065/mcp.2010.0681 | Population |
| Tanenberg 2011 | NA | Population |
| Tanenberg 2013 | 10.1111/papr.12121 | Study design |
| Tasmuth 2002 | NA | Length of follow-up |
| Teixeira 2015 | 10.1590/0004-282X20140232 | Length of follow-up |
| Torre-Mollinedo 2001 | NA | Length of follow-up |
| Tracey 2011 | NA | Study design |
| Treister 2018 | 10.1371/journal.pone.0197844 | Length of follow-up |
| Treves 1991 | NA | Length of follow-up |
| Tuncer 2005 | 10.1163/1568569053421645 | Population |
| Turcotte 2015 | 10.1111/pme.12569 | Intervention |
| Ursini 2011 | http://www.biomedcentral.com/1472-6882/11/46 | Intervention |
| Uzaraga 2012 | NA | Intervention |
| Vadalouca 2010 | NA | Comparator |
| Van Nooten 2015 | NA | Study design |
| Van Seventer 2011 | 10.1186/1477-7525-9-17 | Study design |
| Vanelderen 2015 | 10.1097/ALN.0000000000000508 | Length of follow-up |
| van Seventer 2004 | 10.1111/j.1468-1331.2004.00919.x | No usable data |
| Velasco 2016 | NA | Study design |
| Veldhuijzen 2006 | 10.1177/0269881106061101 | Length of follow-up |
| Venancio-Ramirez 2004 | NA | Length of follow-up |
| Ventafridda 1987 | NA | Comparator |
| Vera-Llonch 2006 | 10.1016/j.ejpain.2005.05.005 | Study design |
| Vidal 2010 | NA | Endpoints |
| Vijayalakshmi 2016 | NA | Length of follow-up |
| Vilming 1986 | 10.1046/j.1468-2982.1986.0603181.x | Length of follow-up |
| Vinik 2014a | 10.2337/dc14-1044 | Length of follow-up |
| Vinik 2014b | NA | Length of follow-up |
| Vinik 2014c | NA | Length of follow-up |
| Vinik 2014d | 10.2337/db14-833-1316 | Duplicate |
| Vinik 2015 | NA | Duplicate |
| Vollmer 2014 | 10.1111/papr.12127 | Length of follow-up |
| Von Delius 2007 | NA | Population |
| Vrethem 1997 | NA | Length of follow-up |
| Wallace 2002 | NA | Population |
| Wallace 2015 | NA | Study design |
| Wang 2017 | 10.3892/etm.2017.4102 | Study design |
| Watson 1982 | 10.1212/WNL.32.6.671 | No usable data |
| Watson 1992 | NA | Length of follow-up |
| Watson 1993 | NA | No usable data |
| Watson 1998 | NA | Length of follow-up |
| Webster 2010 | 10.1111/j1526-4637.2009.00781.x | Intervention |
| Webster 2012 | 10.2147/JPR.S25272 | Intervention |
| Wemicke 2004 | NA | Duplicate |
| Wernicke 2006b | NA | Study design |
| Wernicke 2007 | 10.1111/j.1526-4637.2006.00258.x | Study design |
| Wibbenmeyer 2013 | NA | Population |
| Wibbenmeyer 2014 | 10.1097/BCR.0b013e31828a4828 | Population |
| Wilton 1974 | NA | Intervention |
| Wu 2006 | 10.1016/j.jpain.2006.01.443 | Study design |
| Xuan 2008 | NA | Study design |
| Yarnitsky 2012 | 10.1016/j.pain.2012.02.021 | Study design |
| Yasuda 2016 | NA | Study design |
| Yoshimura 2015 | 10.1053/j.jvca.2015.05.117 | Length of follow-up |
| Young 1985 | 10.1111/j.1464-5491.1985.tb00652.x | Length of follow-up |
| Yousef 2009 | NA | Intervention |
| Yuen 2012 | 10.1002/j.1532-2149.2012.00209.x | Study design |
| Zakerkish 2017 | 10.5812/ircmj.59995 | Length of follow-up |
| Zare 2016 | NA | Population |
| Zarei 2016 | 10.1016/j.clineuro.2016.10.007 | Length of follow-up |
| Zhang 2015 | NA | No usable data |
| Ziegler 2015 | 10.1097/j.pain.0000000000000263 | Length of follow-up |
| Zimmerman 2015 a | NA | Population |
| Zimmerman 2015 b | NA | Population |
| Zis 2016 | NA | Study design |
| Zmarandescu 2013 | 10.1007/s00415-013-6924-0 | No usable data |

### Characteristics of included trials

| Trial | Condition | Trial design | Age | Overall N | N allocated to placebo | N Female | Neuralgia duration (weeks) | Study duration (weeks) | Overall risk of bias | Industry funded | Head to head trial | Drug class |
| --- | --- | --- | --- | --- | --- | --- | --- | --- | --- | --- | --- | --- |
| Allen 2014 | PDPN | RCT, DB, parallel | 60 | 405 | 89 | 108 | 42.1 | 12 | Low | Yes | No | SNRIs |
| Arezzo 2008 | PDPN | RCT, DB, parallel | 58 | 167 | 85 | 64 | 48 | 12 | Low | Yes | No | CC alpha-2-delta ligands |
| Baba 2020 | PDPN | RCT, DB, parallel | 59.8 | 446 | 88 | 158 | 46 | 8 | High | No | No | CC alpha-2-delta ligands |
| Backonja 1998 | PDPN | RCT, DB, parallel | 53 | 165 | 76 | 66 | - | 8 | Low | Yes | No | CC alpha-2-delta ligands |
| Backonja 2008 | PHN | RCT, DB, parallel | 71 | 804 | 196 | 212 | 46.8 | 12 | Low | Yes | No | Capsaicin 8% |
| Clifford 2012 | HIVN | RCT, DB, parallel | 50 | 494 | 162 | 62 | 73.2 | 12 | High | Yes | No | Capsaicin 8% |
| Dworkin 2003 | PHN | RCT, DB, parallel | 72 | 172 | 84 | 92 | 33.8 | 8 | High | Yes | No | CC alpha-2-delta ligands |
| Eerdekens 2016 | PDPN | RCT, DB, parallel | 62 | 127 | 62 | 29 | - | 8 | Low | Yes | No | CC alpha-2-delta ligands |
| Eftekharsadat 2015 | Mixed/other | RCT, DB, parallel | 44 | 90 | 30 | 70 | 21 | 8 | High | No | No | CC alpha-2-delta ligands |
| Freynhagen 2005 | Mixed/other | RCT, DB, parallel | 62 | 338 | 65 | 155 | 37.5 | 8 | Low | Yes | No | CC alpha-2-delta ligands |
| Gao 2010 | PDPN | RCT, DB, parallel | 59 | 215 | 109 | 114 | - | 12 | High | No | No | SNRIs |
| Gao 2015 | PDPN | RCT, DB, parallel | 61 | 405 | 198 | 223 | 39.6 | 12 | Low | Yes | No | SNRIs |
| Goldstein 2005 | PDPN | RCT, DB, parallel | 60 | 457 | 115 | 176 | 44.4 | 12 | High | Yes | No | SNRIs |
| Graff 2000 | PHN | RCT, DB, parallel | 75 | 24 | 11 | 10 | 33.4 | 8 | High | No | No | Tricyclics |
| Guan 2011 | Mixed/other | RCT, DB, parallel | 60 | 308 | 102 | 165 | 25.2 | 8 | High | Yes | No | CC alpha-2-delta ligands |
| Hui 2011 | Mixed/other | RCT, DB, parallel | 52 | 140 | 69 | 214 | - | 8 | Low | Yes | No | CC alpha-2-delta ligands |
| Irving 2011 | PHN | RCT, DB, parallel | 70 | 832 | 408 | 226 | 38.4 | 12 | High | Yes | No | Capsaicin 8% |
| Jang 2017 | Mixed/other | RCT, DB, parallel | 48 | 18 | 9 | 14 | - | 8 | Low | No | No | Tricyclics |
| Jiang 2019 | CIPN | RCT, DB, parallel | 56 | 137 | 69 | 51 | - | 18 | Low | No | No | CC alpha-2-delta ligands |
| Kieburtz 1998 | HIVN | RCT, DB, parallel | 41 | 97 | 50 | 6 | - | 8 | Low | Yes | No | Tricyclics |
| Liu 2014 | Mixed/other | RCT, OL, parallel | 58 | 125 | 33 | 137 | 20 | 12 | High | No | Yes | Tricyclics |
| Liu 2015 | PHN | RCT, DB, parallel | 65 | 219 | 97 | 101 | 4.8 | 8 | Low | Yes | No | CC alpha-2-delta ligands |
| Markman 2018 | Mixed/other | RCT, OL, parallel | 53 | 539 | 208 | - | - | 15 | Low | Yes | No | CC alpha-2-delta ligands |
| Mathieson 2017 | Mixed/other | RCT, DB, parallel | 53 | 209 | 101 | 115 | 2.1 | 8 | Low | No | No | CC alpha-2-delta ligands |
| Moon 2010 | Mixed/other | RCT, DB, parallel | 61 | 240 | 78 | 129 | 40.3 | 8 | Low | Yes | No | CC alpha-2-delta ligands |
| Mu 2017 | PDPN | RCT, DB, parallel | 61 | 620 | 307 | 327 | 27.6 | 8 | Low | Yes | No | CC alpha-2-delta ligands |
| NCT 00394901 | PHN | RCT, DB, parallel | 70 | 369 | 97 | 173 | - | 12 | Low | Yes | No | CC alpha-2-delta ligands |
| Raskin 2005 | PDPN | RCT, DB, parallel | 59 | 348 | 113 | 186 | 51.6 | 12 | High | Yes | No | SNRIs |
| Rauck 2012 | PDPN | RCT, DB, parallel | 59 | 420 | 120 | 249 | - | 12 | Low | Yes | Yes | CC alpha-2-delta ligands |
| Rosenstock 2004 | PDPN | RCT, DB, parallel | 60 | 146 | 69 | 64 | - | 8 | High | Yes | No | CC alpha-2-delta ligands |
| Rowbotham 1998 | PHN | RCT, DB, parallel | 74 | 225 | 116 | 107 | - | 8 | High | Yes | No | CC alpha-2-delta ligands |
| Sabatowski 2004 | PHN | RCT, DB, parallel | 72 | 238 | 81 | 131 | 31 | 8 | High | Yes | No | CC alpha-2-delta ligands |
| Sang 2013 | PHN | RCT, DB, parallel | 66 | 450 | 230 | 283 | - | 10 | Low | Yes | No | CC alpha-2-delta ligands |
| Satoh 2010 | PDPN | RCT, DB, parallel | 61 | 314 | 135 | 77 | 38.4 | 12 | High | Yes | No | CC alpha-2-delta ligands |
| Serpell 2002 | Mixed/other | RCT, DB, parallel | 57 | 305 | 152 | 164 | 57.6 | 8 | High | Yes | No | CC alpha-2-delta ligands |
| Shlay 1998 | HIVN | RCT, DB, parallel | - | 136 | 53 | - | - | 14 | Low | No | No | Tricyclics |
| Simpson 2001 | PDPN | RCT, DB, parallel | 50 | 60 | 27 | 24 | - | 8 | High | No | No | CC alpha-2-delta ligands |
| Simpson 2008 | HIVN | RCT, DB, parallel | 48 | 307 | 82 | 21 | 58.8 | 12 | Low | Yes | No | Capsaicin 8% |
| Simpson 2010 | HIVN | RCT, DB, parallel | 48 | 302 | 147 | 57 | 73.2 | 14 | Low | Yes | No | CC alpha-2-delta ligands |
| Simpson 2014 | HIVN | RCT, DB, parallel | 47 | 375 | 192 | 114 | 28.8 | 16 | Low | Yes | No | CC alpha-2-delta ligands |
| Simpson 2017 | PDPN | RCT, DB, parallel | 63 | 369 | 183 | 154 | 69.6 | 12 | Low | Yes | No | Capsaicin 8% |
| Tolle 2008 | PDPN | RCT, DB, parallel | 59 | 395 | 93 | 176 | - | 12 | Low | Yes | No | CC alpha-2-delta ligands |
| van Seventer 2006 | PHN | RCT, DB, parallel | 71 | 366 | 93 | 200 | 27 | 12 | High | Yes | No | CC alpha-2-delta ligands |
| van Seventer 2010 | Mixed/other | RCT, DB, parallel | 51 | 254 | 127 | 129 | 52.8 | 8 | Low | Yes | No | CC alpha-2-delta ligands |
| Wallace 2010 | PHN | RCT, DB, parallel | 67 | 400 | 131 | 199 | - | 10 | Low | Yes | No | CC alpha-2-delta ligands |
| Webster 2010a | PHN | RCT, DB, parallel | 71 | 598 | 77 | 149 | 45.6 | 12 | High | Yes | No | Capsaicin 8% |
| Webster 2010b | PHN | RCT, DB, parallel | 70 | 310 | 106 | 83 | 36 | 12 | Low | Yes | No | Capsaicin 8% |
| Wernicke 2006 | PDPN | RCT, DB, parallel | 61 | 334 | 106 | 130 | 45.6 | 12 | High | Yes | No | SNRIs |
| Yasuda 2011 | PDPN | RCT, DB, parallel | 61 | 339 | 167 | 82 | 51.6 | 12 | Low | Yes | No | SNRIs |
| Zhang 2013 | PHN | RCT, DB, parallel | 62 | 371 | 95 | 182 | - | 12 | Low | Yes | No | CC alpha-2-delta ligands |

CC, calcium channel. DB, double blind. HIVN, Human Immunodeficiency Virus-induced neuropathy. N, number. OL, open label. PDPN, painful diabetic peripheral neuropathy. PHN, post-herpetic neuralgia. RCT, randomized controlled trial. SNRI, serotonin-norepinephrine reuptake inhibitor.

### Risk of bias of included trials

| Trial | Random sequence generation | Allocation concealment | Performance bias | Detection bias | Attrition bias | Selective reporting bias | Other | Overall bias rating |
| --- | --- | --- | --- | --- | --- | --- | --- | --- |
| Allen 2014 | Unclear | Low | Low | Low | Unclear | Low | Low | Low |
| Arezzo 2008 | Low | Low | Low | Low | Unclear | Low | Low | Low |
| Baba 2020 | Unclear | Low | Low | Unclear | High | Low | Low | High |
| Backonja 1998 | Low | Low | Low | Low | Low | Unclear | Low | Low |
| Backonja 2008 | Unclear | Low | Low | Low | Low | Unclear | Low | Low |
| Clifford 2012 | Unclear | Unclear | Unclear | Unclear | Low | High | Low | High |
| Dworkin 2003 | Unclear | Low | Low | Low | High | Unclear | Low | High |
| Eerdekens 2016 | Unclear | Low | Low | Low | Unclear | Low | Low | Low |
| Eftekharsadat 2015 | Unclear | Low | High | Unclear | Unclear | Low | Low | High |
| Freynhagen 2005 | Unclear | Low | Low | Low | Unclear | Low | Low | Low |
| Gao 2010 | Unclear | Unclear | Low | Low | Unclear | Unclear | Low | High |
| Gao 2015 | Unclear | Low | Low | Low | Low | Low | Low | Low |
| Goldstein 2005 | Unclear | Low | High | High | Low | Unclear | Low | High |
| Graff 2000 | Unclear | Unclear | Unclear | Unclear | Low | Unclear | Low | High |
| Guan 2011 | Unclear | Unclear | Low | Low | Unclear | Low | Low | High |
| Hui 2011 | Low | Low | Low | Low | Low | Low | Low | Low |
| Irving 2011 | Unclear | Unclear | Low | Low | Low | Unclear | Low | High |
| Jang 2017 | Unclear | Unclear | Low | Low | Low | Low | Low | Low |
| Jiang 2019 | Unclear | Low | Low | Low | Low | Low | Low | Low |
| Kieburtz 1998 | Unclear | Low | Low | Low | Low | Unclear | Low | Low |
| Liu 2014 | Unclear | Unclear | High | High | Unclear | Unclear | Low | High |
| Liu 2015 | Low | Low | Low | Low | Low | Low | Low | Low |
| Markman 2018 | Low | Low | Unclear | Unclear | Low | Low | Low | Low |
| Mathieson 2017 | Low | Low | Low | Low | Low | Low | Low | Low |
| Moon 2010 | Low | Low | Low | Low | Unclear | Low | Low | Low |
| Mu 2017 | Low | Low | Low | Low | Unclear | Unclear | Low | Low |
| NCT 00394901 | Unclear | Low | Unclear | Low | Low | Low | Low | Low |
| Raskin 2005 | Unclear | Low | Low | Low | Unclear | Unclear | Low | High |
| Rauck 2012 | Low | Low | Low | Low | Low | Low | Low | Low |
| Rosenstock 2004 | Low | Low | Unclear | Unclear | Unclear | Unclear | Low | High |
| Rowbotham 1998 | Unclear | Low | Low | Low | Unclear | Unclear | Low | High |
| Sabatowski 2004b | Low | Unclear | Low | Low | Unclear | Unclear | Low | High |
| Sang 2013 | Low | Unclear | Low | Low | Low | Low | Low | Low |
| Satoh 2010 | Low | Unclear | Unclear | Unclear | Unclear | Low | Low | High |
| Serpell 2002 | Low | Low | Unclear | Unclear | Unclear | Unclear | Low | High |
| Shlay 1998 | Unclear | Low | Low | Low | Low | Low | Low | Low |
| Simpson 2001 | Unclear | Unclear | Low | Low | Unclear | Unclear | Low | High |
| Simpson 2008 | Unclear | Unclear | Low | Low | Low | Low | Low | Low |
| Simpson 2010 | Unclear | Low | Low | Low | Unclear | Low | Low | Low |
| Simpson 2014 | Unclear | Low | Low | Low | Low | Low | Low | Low |
| Simpson 2017 | Low | Low | Low | Low | Unclear | Low | Low | Low |
| Tolle 2008 | Unclear | Low | Low | Low | Low | Unclear | Low | Low |
| van Seventer 2006 | Unclear | Unclear | Unclear | Unclear | Unclear | High | Low | High |
| van Seventer 2010 | Unclear | Low | Low | Unclear | Low | Low | Low | Low |
| Wallace 2010 | Unclear | Low | Low | Low | Low | Low | Low | Low |
| Webster 2010a | Unclear | Unclear | Low | Low | Low | Unclear | Low | High |
| Webster 2010b | Unclear | Unclear | Low | Low | Low | Low | Low | Low |
| Webster 2011 | Low | Low | High | High | Low | Low | Low | High |
| Wernicke 2006 | Unclear | Low | Low | Unclear | Unclear | Unclear | Low | High |
| Yasuda 2011 | Low | Low | Low | Unclear | Low | Low | Low | Low |
| Zhang 2013 | Low | Low | Low | Low | Unclear | Unclear | Low | Low |

### Forest plots

#### PIR≥50%

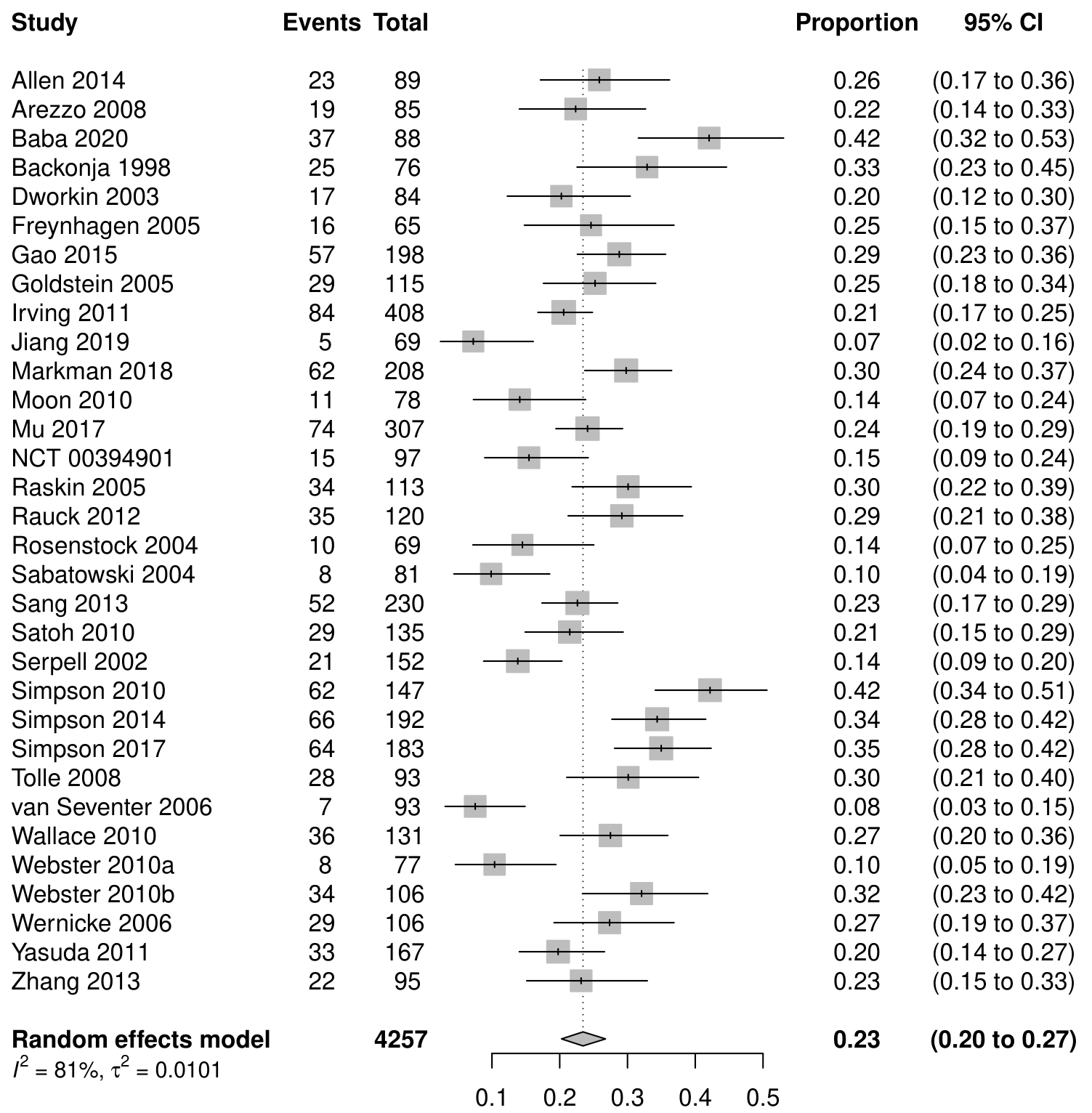

#### PGIC much or very much improved

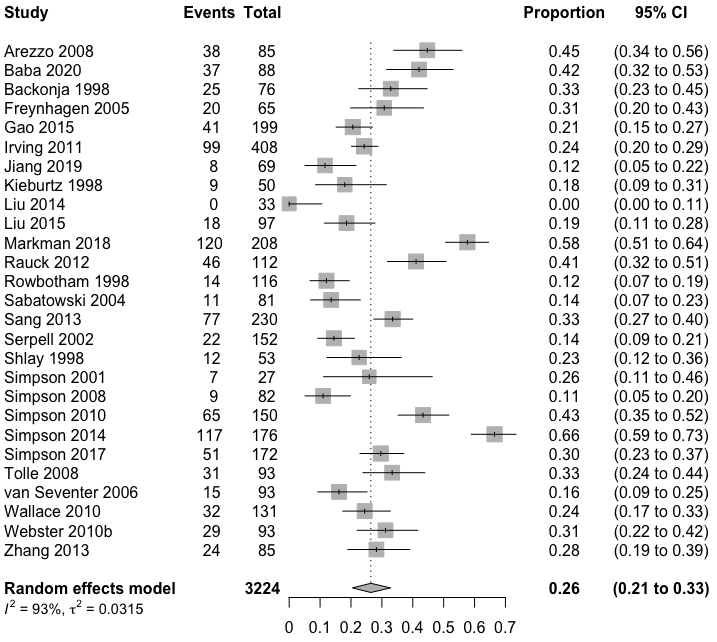

#### Serious adverse events

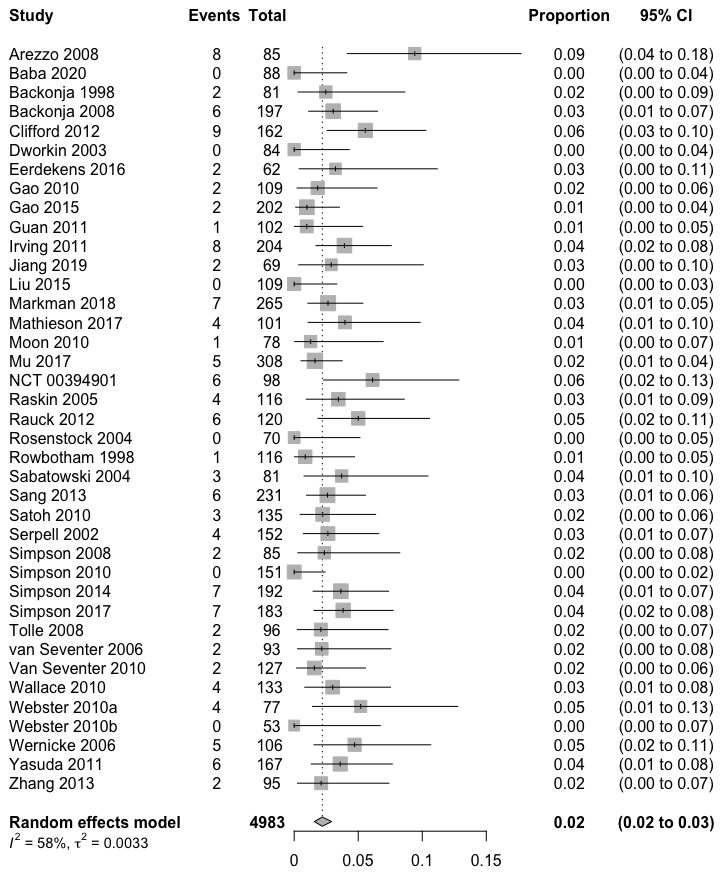

#### Discontinuations due to adverse events

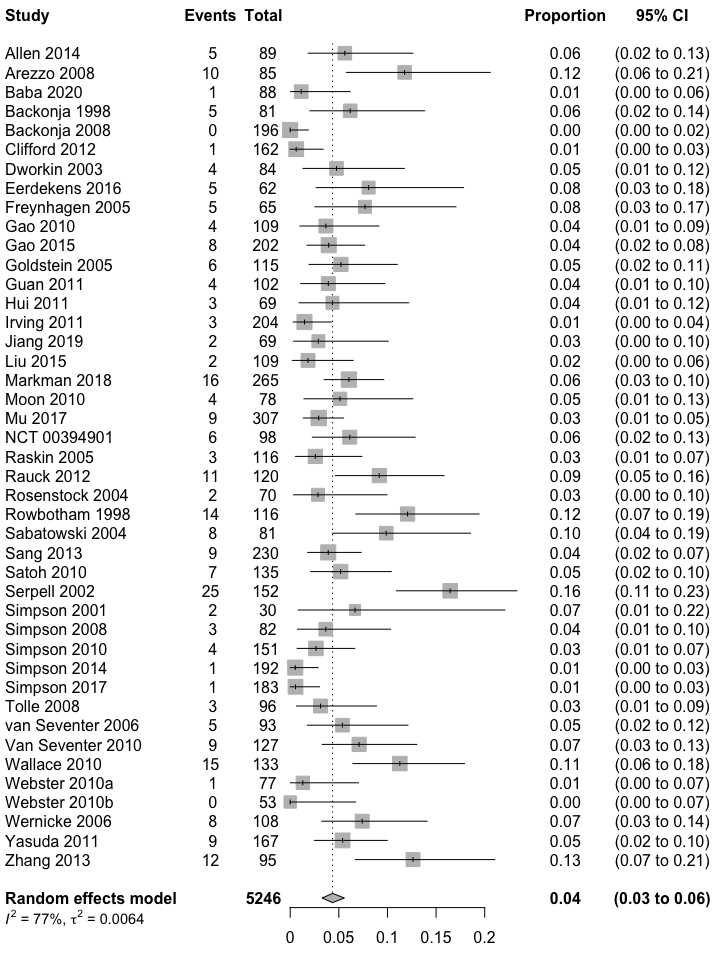

#### Discontinuations due to lack of efficacy

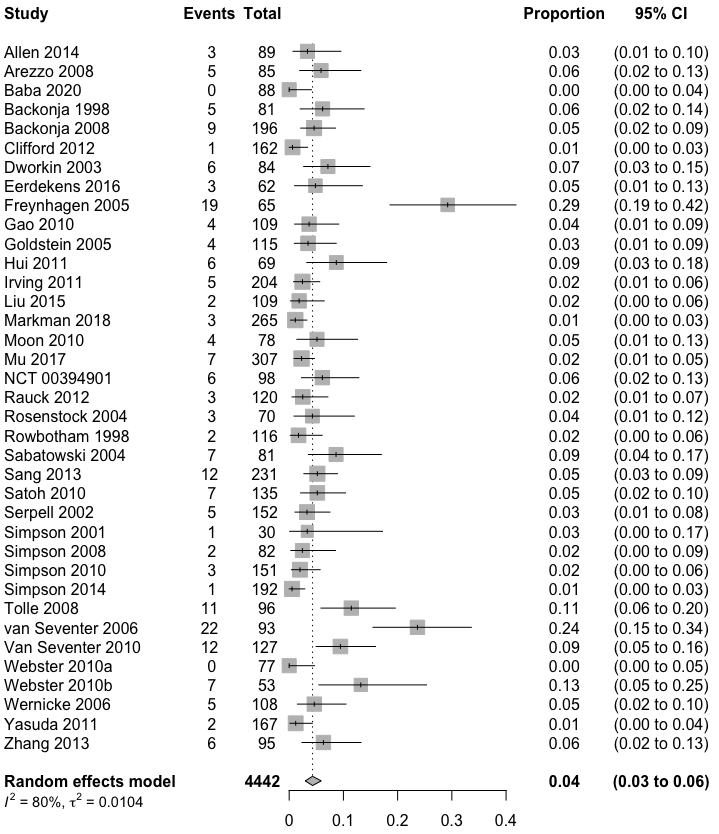

### Multicollinearity

#### Graphical display for PIR ≥30%

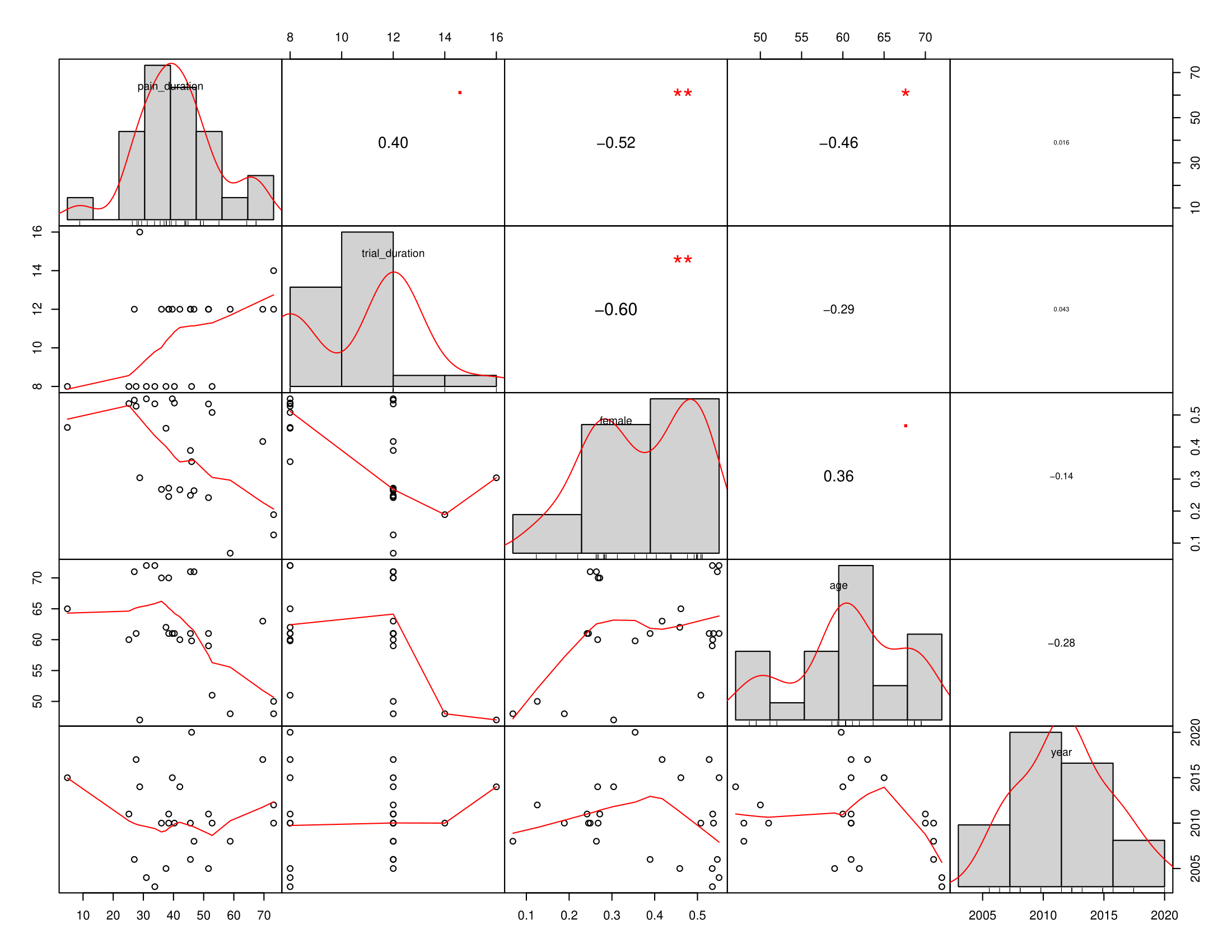

#### Graphical display for adverse events

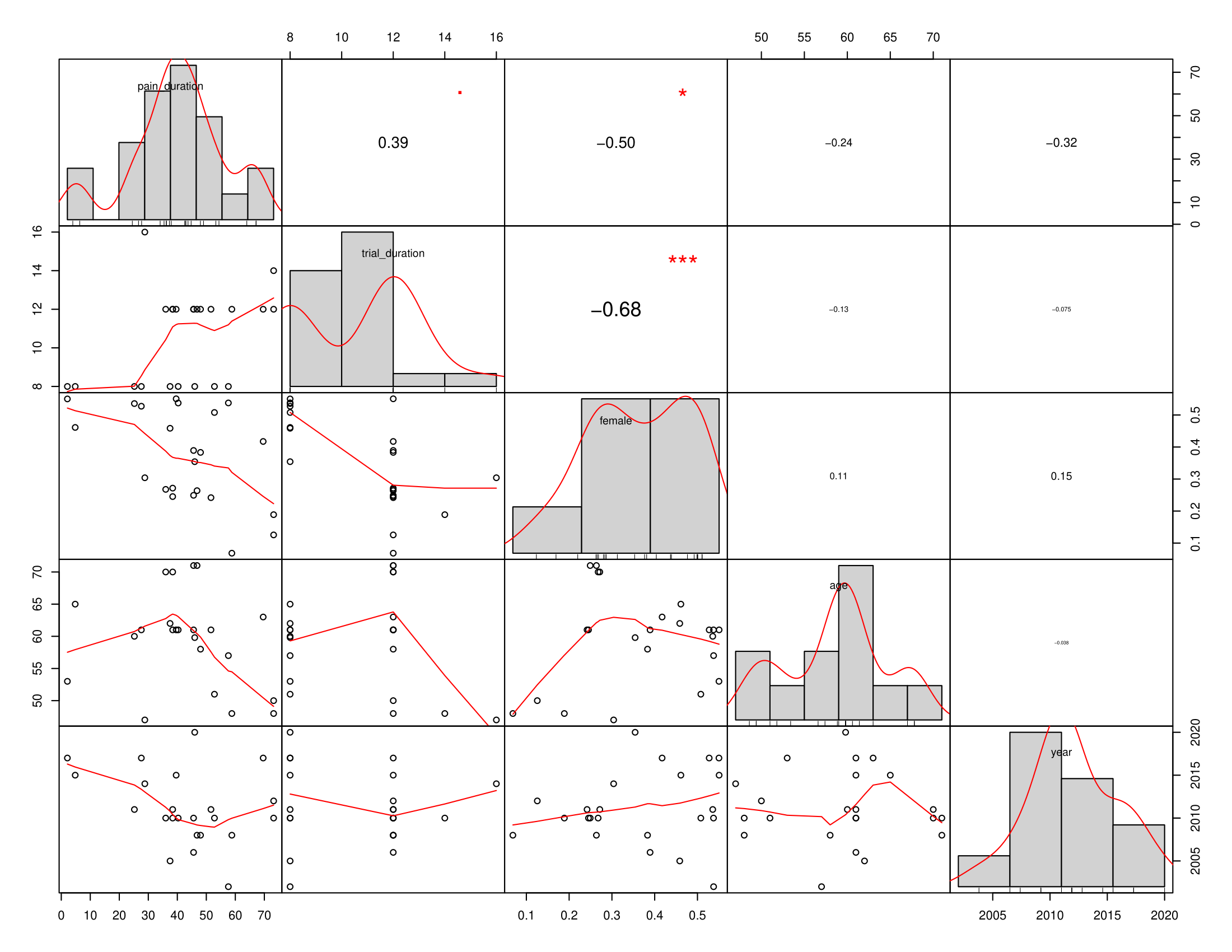
